# Bacterial community modifies host genetics effect on early childhood caries

**DOI:** 10.1101/2023.01.11.23284235

**Authors:** Freida Blostein, Tianyu Zou, Deesha Bhaumik, Elizabeth Salzman, Kelly M. Bakulski, John R. Shaffer, Mary L. Marazita, Betsy Foxman

## Abstract

**Background:** By age five approximately one-fifth of children have early childhood caries (ECC). Both the oral microbiome and host genetics are thought to influence susceptibility. Whether the oral microbiome modifies genetic susceptibility to ECC has not been tested. We test whether the salivary bacteriome modifies the association of a polygenic score (PGS, a score derived from genomic data that summarizes genetic susceptibility to disease) for primary tooth decay on ECC in the Center for Oral Health Research in Appalachia 2 longitudinal birth cohort.

**Methods:** Children were genotyped using the Illumina Multi-Ethnic Genotyping Array and underwent annual dental examinations. We constructed a PGS for primary tooth decay using weights from an independent, genome-wide association meta-analysis. Using Poisson regression, we tested for associations between the PGS (high versus low) and ECC incidence, adjusting for demographic characteristics (n=783). An incidence-density sampled subset of the cohort (n=138) had salivary bacteriome data at 24- months of age. We tested for effect modification of the PGS on ECC case status by salivary bacterial community state type (CST).

**Results:** By 60-months, 20.69% of children had ECC. High PGS was not associated with an increased rate of ECC (incidence-rate ratio:1.09 (95% confidence interval (CI): 0.83, 1.42)). However, having a cariogenic salivary bacterial CST at 24-months was associated with ECC (odds ratio (OR): 7.48 (95%CI: 3.06, 18.26)), which was robust to PGS adjustment. An interaction existed between the salivary bacterial CST and the PGS on the multiplicative scale (P= 0.04). The PGS was associated with ECC (OR: 4.83 (95% CI: 1.29, 18.17)) only among individuals with a noncariogenic salivary bacterial CST (n=70).

**Conclusions:** Genetic causes of caries may be harder to detect when not accounting for cariogenic oral microbiomes. As certain salivary bacterial CSTs increased ECC-risk across genetic-risk strata, preventing colonization of cariogenic microbiomes would be universally beneficial.

## Background

In 2016, 20% of US children aged 2-5 years had early childhood caries (ECC): one or more lesion in a primary tooth (Fleming and Afful 2018). ECC is associated with low-self-esteem, disrupted language acquisition, and poor oral health throughout the life course (Martins et al. 2017). Dental caries is a multifactorial disease, influenced by genetic, microbial, and sociodemographic factors, wherein teeth are demineralized by acidic byproducts of oral microbes (Pitts et al. 2017).

Dental caries prevalence clusters by family, and genetic causes for dental caries are biologically plausible (Boraas et al. 1988; Haworth et al. 2018; Shungin et al. 2019). Yet heritability estimates of caries vary across populations, and genome-wide association studies (GWAS) have reported few significant and reproducible loci (Boraas et al. 1988; Bretz et al. 2005; Wang et al. 2010; Shaffer, Feingold, et al. 2012; Shaffer, Wang, et al. 2012; Zeng et al. 2014; Shaffer et al. 2015; Haworth et al. 2018; Shungin et al. 2019; Haworth et al. 2020). This suggests a polygenic architecture, wherein many genetic variants cumulatively contribute small effects (Mayhew and Meyre 2017).

Previous studies of cumulative genetic risk in dental decay have tested for joint associations between a few candidate genes or single nucleotide polymorphisms (SNPs) (Wang et al. 2012; Bezamat et al. 2021). An alternative to candidate gene approaches is to instead create a polygenic score (PGS). With PGSs, SNPs associated with a phenotype in a previous GWAS are summed together in an independent population, creating a summary score. This score can be weighted by the previous GWAS’s effect estimates (Lewis and Vassos 2020). Although PGS have been successfully applied in other phenotypes, we found no study using a PGS for ECC (Lewis and Vassos 2020).

Environmental risk factors, such as the oral microbiome, may also modify genetic risk for ECC, creating variance in heritability across populations (Mayhew and Meyre 2017). Previously, the acidogenic bacteria *Streptococcus mutans* was considered the primary microbial agent of ECC (Pitts et al. 2017). However, the salivary microbiome of children under two years of age can predict future ECC with reasonable accuracy, before salivary *S. mutans* detection (Grier et al. 2020; Blostein et al. 2022), suggesting an ecological dysbiosis in the microbiome contributes to ECC (Takahashi and Nyvad 2010; Pitts et al. 2017). Prior studies testing for interactions between bacterial and host genetic risk factors for ECC have considered only *S. mutans* (Slayton et al. 2005; Meng et al. 2019). We found no analyses of interactions between host genetics and bacterial communities in ECC.

In this analysis, we explored gene-environment interactions in ECC using a PGS and salivary bacterial communities. We constructed a PGS and tested for associations with ECC in a longitudinal birth cohort of 783 children. Using a nested case-control subset of the cohort with 16S rRNA amplicon sequencing data from saliva samples (n=138), we tested if the effect of the PGS was modified by the salivary bacterial community at 24-months of age.

## Methods

### Study cohort

We analyzed data from the Center for Oral Health Research in Appalachia 2 study (COHRA2). COHRA2 recruited white, pregnant women between 2011 and 2018 from Pennsylvania and West Virginia (Supplemental Methods) (Neiswanger et al. 2015). The study has IRB approval from the University of Pittsburgh and West Virginia University. All potential participants had the study explained to them in detail and the women signed consent forms prior to research assessments. This analysis adheres to the Strengthening the Reporting of Observational studies in Epidemiology guidelines.

Mothers attended one prenatal in-person visit at enrollment. Mothers and children attended in-person visits when the child was approximately 2-, 12-, 24-, 36-, 48- and 60-months of age. Pennsylvanian participants also attended visits at the child’s birth and first primary tooth emergence. For this analysis, we selected child-visits up to and including the 60-month visit (2022 data freeze). At visits, trained and calibrated dental professionals conducted a comprehensive dental examination (Neiswanger et al. 2015). Caries assessment was performed using the PhenX Toolkit Dental Caries Experience Prevalence Protocol (http://www.phenxtoolkit.org/, protocol number 080300). The examination included collection of salivary microbial samples in OMNIgene Discover kits (OM-501/505 DNA Genotek) and human DNA in Oragene kits. Kits were stored at -80-degrees-Celsius.

### Early childhood caries

We defined ECC as any white spots, lesions, or fillings in children’s primary dentition at or before their 60-month visit. To assess phenotype severity, we also considered the count of decayed (including white spots) or filled primary tooth surfaces, and an ECC definition excluding white spots.

### Human genetic variables

Children’s samples were genotyped using the Illumina Multi-Ethnic Global-8 v1.0 Array (∼1.7 million SNPs). We excluded participants with high or low heterozygosity (n=5) or mismatches between reported and genetic sex (n=9). We removed ambiguous or duplicated SNPs and SNPs with minor allele frequency≤0.01, deviation from Hardy-Weinberg equilibrium based on P- value<1* 10^-6^, or genotyping rate<0.1. Ungenotyped SNPs and sporadicly missing genotyped SNPs were imputed using the Michigan Imputation Server (Minimac 3 for autosomal chromosomes, and Minimac 4 for chromosome X), and Haplotype Reference Consortium phase 1 as the reference panel (Das et al. 2016; the Haplotype Reference Consortium 2016).

Genetic ancestry principal components (PCs) and PGSs for ECC were calculated using PLINK v1.9. PCs were z-score standardized. For PGS weights, we used effect estimates from a GWAS meta-analysis of 22 European-ancestry cohorts (base data, n=17,666 children) (Haworth et al. 2018). After quality-control, 6,044,259 SNPs overlapped between the base data and COHRA2 (Supplemental Methods). PGSs were calculated at 7 P- value thresholds: 0.001, 0.05, 0.1, 0.2, 0.3, 0.4 and 0.5. We selected the PGS with the best pseudo-R^2^ measures (Lee et al. 2012) in cross-sectional models regressing prevalent ECC against each PGS, child sex, and the first 5 genetic ancestry PCs (Supplemental Methods). For SNPs in the PGS, we provide the identifier, chromosome, position, weight, and P-value from the base data (doi:10.6084/m9.figshare.21890670), consistent with reporting standards (Haworth et al. 2018; Wand et al. 2021). To facilitate interpretation, the PGS was z-score standardized and dichotomized into high (≥0) and low PGS (<0) for some analyses.

### Nested case-control study

To incorporate the salivary microbiome, we performed a secondary analysis of a previously conducted incidence-density sampled case-control study nested within COHRA2 (2019 data freeze) (Blostein et al. 2022). In this substudy, cases were children with primary dentition white spots, dental lesions, or fillings at or before the 60-month visit. For visits at which cases were diagnosed, an approximately equivalent number of children without dental decay were sampled as age-at-incidence-matched controls (Appendix Figure 1). For cases and controls, banked saliva samples from the visit of case diagnosis and preceding visits were selected for 16S rRNA amplicon sequencing. Sequences were processed using DADA2 (Supplemental Methods).

#### Salivary bacterial measures

In the initial case-control analysis (Blostein et al. 2022), we identified samples with similar bacterial communities (community state types (CSTs)) using Dirichlet multinomial mixture models (Holmes et al. 2012). Two CSTs characterized by *Streptococcus* predominated at the 2-month visit and were not associated with case status. At later visits, two CSTs characterized by *Haemophilus parainfluenzae* and *Nesisseria* were more common among controls and two CSTs characterized by *Streptococcus* and *Veillonella* were more common among cases (Blostein et al. 2022). To preserve sample size within CST-strata, we collapsed together the two 2-month visit CSTs (‘Early-life *Streptococcus*-dominated CSTs’), the two control-associated CSTs (*Haemophilus/Neisseria* CSTs), and the two case-associated CSTs (*Streptococcus/Veillonella* CSTs). In the initial analysis (Blostein et al. 2022), case-control differences were apparent at the 12- and 24-month visits, so we used those visits in this secondary analysis.

We used other bacterial variables, such as salivary *Streptococcus mutans* presence and the Chao1 and Shannon alpha diversity indexes in sensitivity analyses (Supplemental Methods).

#### Other variables

We identified confounders and precision variables through literature review.

The following variables were ascertained from maternal interview: maternal-reported child race (white versus bi/multi-racial), child ethnicity (Latino/Hispanic versus not Latino/Hispanic), child sex (male versus female), maternal education at prenatal visit (greater than high school versus high school degree or less), prenatal maternal illness, prenatal maternal antibiotics, and prenatal maternal smoking.

The following variables were identified as potential microbiome-ECC confounders: count of primary teeth (dental exam), child antibiotics within 3 months of visit, sugar-sweetened beverage consumption, breastfeeding, and delivery method (vaginal versus C-section) (Supplemental Methods).

### Statistical analyses

All statistical analyses were conducted using R version 4.0.2.

#### Exclusion criteria

Before calculating genetic ancestry PCs, we selected single representatives from among related children (siblings and cousins, n=26 children excluded) and excluded any children not reported as white by their mothers (n=238). Although race and genetic ancestry are not synonymous, child race was highly concordant with genetic ancestry (Appendix Figure 2). We excluded children missing covariates, visualizing exclusions as a CONSORT diagram (Appendix Figure 3).

#### Descriptive statistics

We used Welch’s two sample t-test (continuous variables) and Pearson’s Chi-squared test (categorical variables) to test for differences by ever ECC and by dichotomized PGS. To investigate the impact of age-at-diagnosis, we tested for differences by prevalent ECC within visit-strata. In the nested case-control substudy, we tested for differences by CST at the 12- and 24-month visits.

We visualized the PGS distribution by categorical variables using violin with captured box plots. We visualized correlation between continuous variables and the PGS using scatter plots.

#### Regression analyses: entire cohort

We calculated ECC incidence rate ratios (IRRs) for the dichotomized PGS among the entire cohort using Poisson regression with child age at diagnosis/censoring as an offset and the robust variance estimate. We performed the following adjustments: base model (first 5 genetic ancestry PCs), demographic model (base model+child sex+maternal education), and health behaviors model (demographic model+prenatal maternal exposures (antibiotic use, illness, smoking)+breastfeeding duration).

As a sensitivity analysis to investigate the impact of age-at-diagnosis, we modeled ECC prevalence ratios (PRs) at the 36-, 48-, and 60-month-visits using modified Poisson regression (Zou 2004).

As a sensitivity analysis to investigate loss-to-follow-up, we used inverse probability weights (Supplemental Methods).

#### Effect modification analyses: case-control subset

We calculated odds ratios (ORs) in the case-control substudy using logistic regression (Supplemental Methods). We regressed case status on the dichotomized PGS, controlling for demographic variables and visit of matching. Next, we included the 24-month CST and microbiome-ECC confounders. To evaluate effect-modification on the multiplicative scale, we included a CST*PGS interaction. We reported strata-specific ORs (Knol and VanderWeele 2012). We calculated additive interaction measures using the interactionR package and MOVER method for estimating confidence intervals (Zou 2008; Alli 2021).

We visualized effect modification using a forest plot and plotted the proportion of cases by PGS quartile and CST. To visualize the joint effect on disease progression, we plotted the number of decayed and filled tooth surfaces (including white spots) at the 60-month visit by CST and PGS among cases.

As sensitivity analyses, we also used 12-month CST or 24-month *S. mutans* presence rather than 24-month CST as the effect modifier.

## Results

### Study descriptive statistics in entire cohort

Of the 1131 COHRA2 children, 783 had complete covariate data (Appendix Figure 3, Appendix Table 1). Of these, 162 had ECC by the 60-month visit. The earliest diagnosis was at the 12-month visit, but most diagnoses occurred at or after the 36-month visit (Appendix Table 2).

### PGS not associated with ECC in bivariate analyses in entire cohort

ECC was associated with maternal education, prenatal smoking, and breastfeeding duration (Table **1**).

**Table 1:**
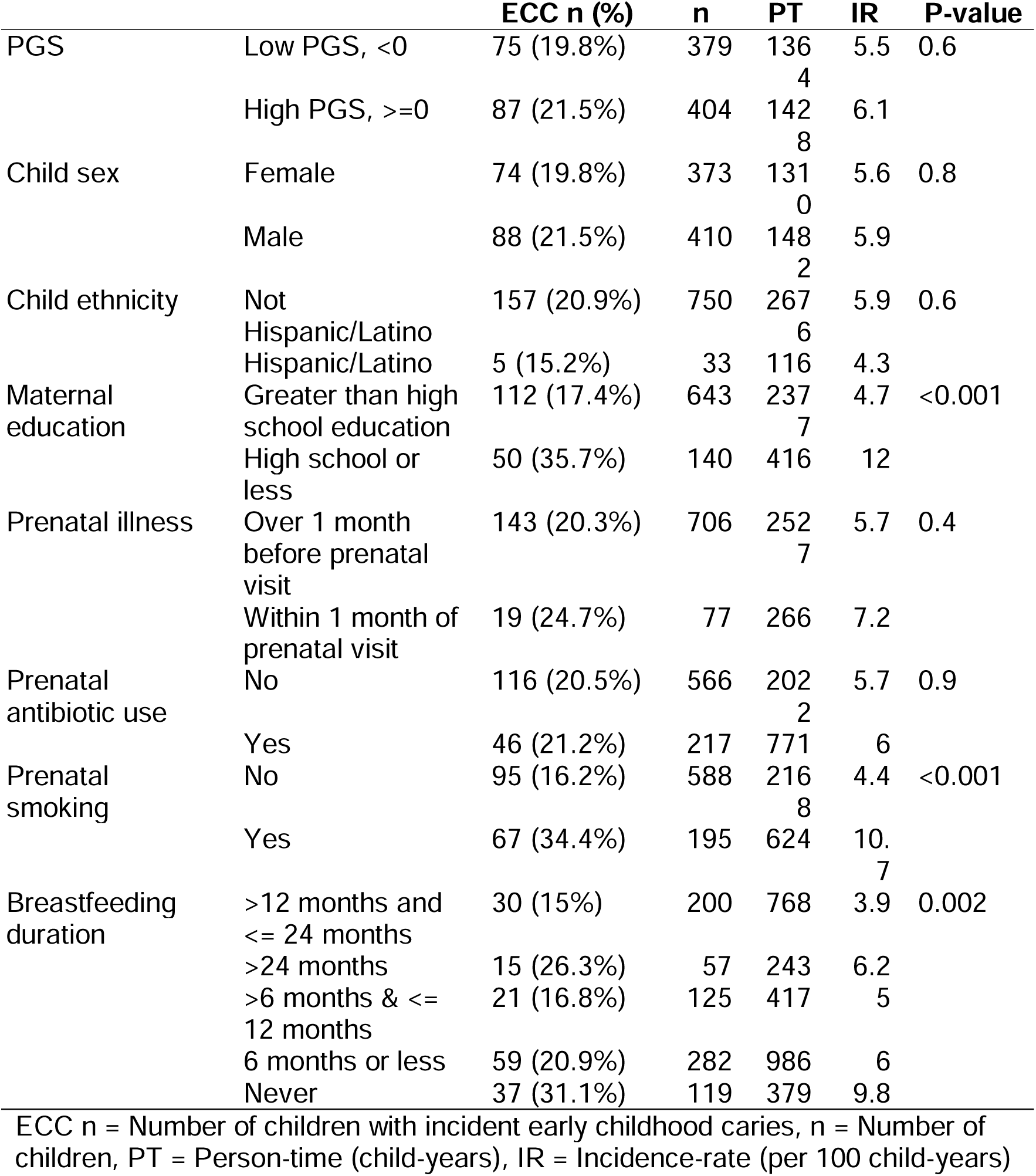
Cumulative incidence and incidence rate of first diagnosis of early childhood caries (ECC) by polygenic score (PGS) and other covariates among 783 Appalachian children

Based on *R*^2^ metrics, we selected the PGS for ECC constructed using a *P*- value threshold=0.001 (Appendix Figures 4 & 5). The PGS was not significantly associated with ECC (ever ECC mean PGS=0.06 versus never ECC mean PGS=0.00, *P*-value=0.5). There was some variability in the PGS by visit-of-diagnosis (Figure 1). Results were similar when excluding white spots when defining ECC (Appendix Figure 6).

**Figure 1:**
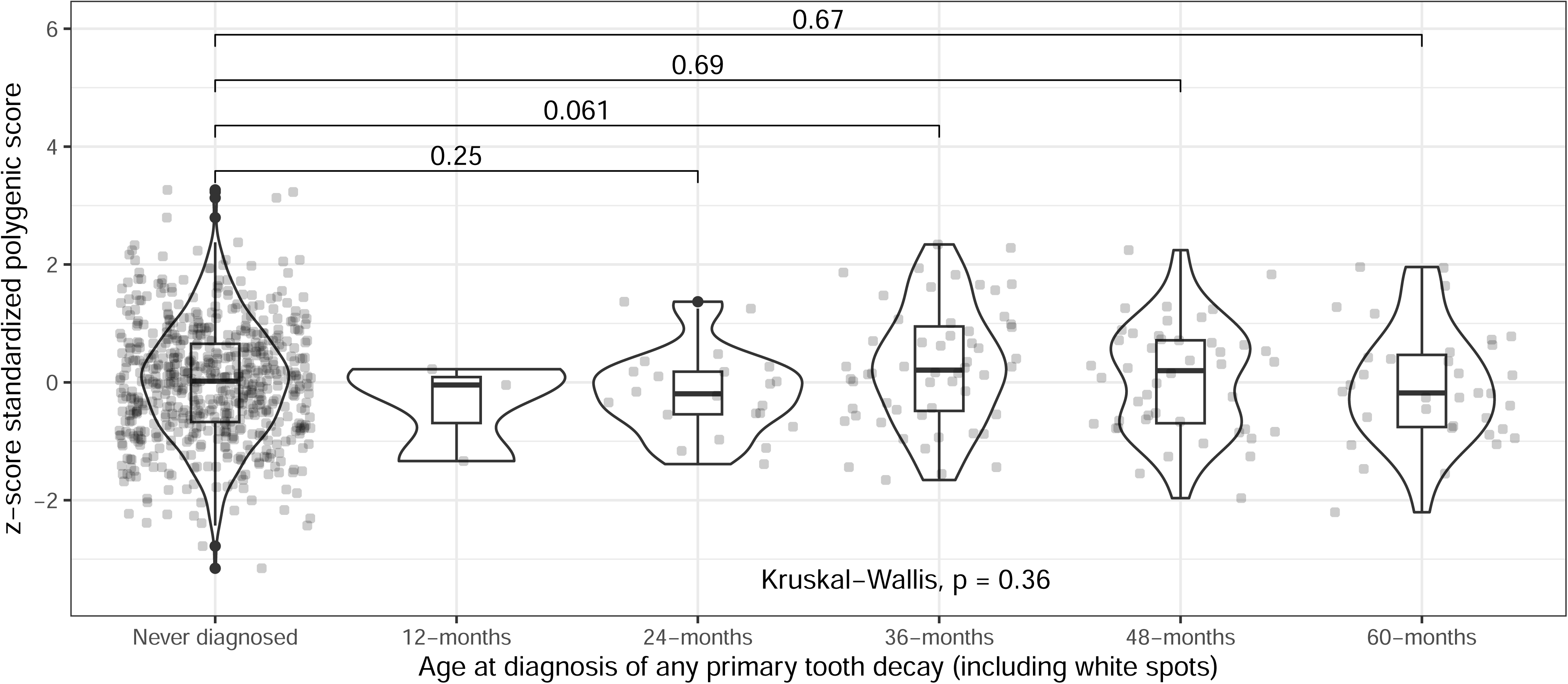
Distributions of z-score standardized polygenic scores by age of first early childhood caries diagnosis among 783 Appalachian children in a longitudinal cohort

Children with a high versus low PGS did not differ in any tested variables (Appendix Table 3).

### PGS not significantly associated with ECC incidence in multivariable models among the entire cohort

A high PGS was not associated with ECC incidence in multivariable models (demographic model IRR=1.09 (95% CI: 0.83,1.42), Table **2**).

**Table 2:** Results from Poisson regression models of incident early childhood caries (ECC) among 783 Appalachia children followed from birth until up to 5- years of age. Robust standard errors reported in confidence intervals.

|  | **IRR** | **95% CI** | **P val** |
| --- | --- | --- | --- |
| Base model | 1.09 | 0.83, 1.44 | 0.53 |
| Demographic model | 1.09 | 0.83, 1.42 | 0.55 |
| Health behaviors model | 1.07 | 0.82, 1.4 | 0.61 |

Base model: z-score standardized genetic ancestry principal components, Demographic model: base model + child sex + maternal education, Health behaviors model: demographic model + prenatal illness + prenatal antibiotics + prenatal smoking + breastfeeding duration (years), IRR: incidence rate ratio, CI: 95% confidence interval

There was a stronger association between prevalent ECC and the dichotomized PGS at the 36-month visit (PR:1.55 (95%CI: 0.99,2.42),*P*=0.16) than at the 48-month (PR:1.41 (95%CI:1.01,1.96),*P*=0.05) or 60-month visit (PR:1.26 (95%CI:0.91,1.74),*P*=0.24, Appendix Table 4).

Inverse probability weighted effect estimates were similar but slightly attenuated (Supplemental Results).

### Study descriptive statistics in nested case-cohort subset

The case-control subset was similar to the entire cohort. However, since the case-control design oversampled cases, a greater proportion of mothers in the case-control subset reported a high school education or less and prenatal smoking (Appendix Table 5).

### Salivary CST was associated with ECC in bivariate analyses in the case-control subset

Case status was not significantly associated with PGS (mean case PGS=0.13 versus mean control PGS=0.05, *P*-value=0.6), but was associated with maternal education and prenatal smoking (Appendix Table 6).

Controls were more likely than cases to have a *Haemophilus/Neisseria* CST at both 12-months (25 (36%) cases versus 52 (80%) controls *P*-value<0.001) and 24-months (21 (29%) cases versus 49 (74%) controls *P*-value<0.001).

Cases (24 (33%)) were more likely than controls (2 (3.0%), *P*-value<0.001) to have *Streptococcus mutans* at 24-months, but not 12-months (4 (5.8%) cases versus 1 (1.5%) control, *P*-value=0.4).

Cases had higher sugar-sweetened beverage consumption than controls (Appendix Table 7). Children with a *Haemophilus/Neisseria* CST had lower sugar-sweetened beverage consumption than children with a *Streptococcus/Veillonella* CST at 24-months (Appendix Figure 7).

The PGS was not associated with alpha diversity, CST, or *S. mutans* detection (Appendix Figures 7 & 8).

### PGS and bacterial CST display antagonistic interaction in the nested case-control subset

In the nested case-control subset, as in the entire cohort, there was no significant association between the PGS and ECC (Appendix Table 8, Figure **2**A).

**Figure 2:**
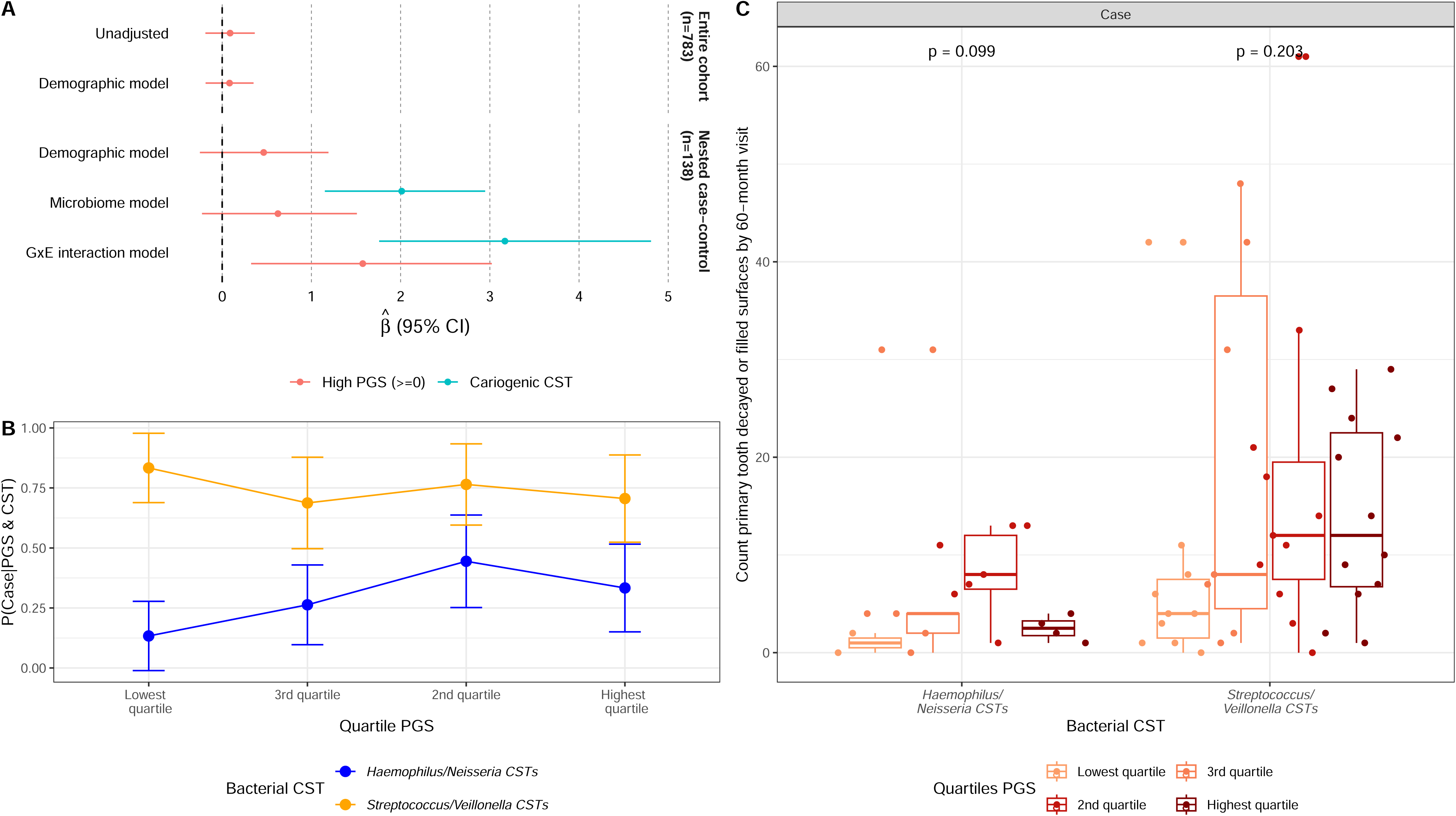
Results from effect modification analysis for effect of polygenic score (PGS) on early childhood caries by salivary bacterial community state type (CST) among Appalachian children A) Regression results in entire cohort (n=783, top facet) and in incidence-density sample subset (n=138, bottom facet). B) Proportion of cases (y-axis) among individuals in each quartile of PGS (x-axis) and with each CST (colors) (n=138). C) Count of decayed and filled primary tooth surfaces (including white spots) at 60-month visit among all cases in nested case-control subset by quartile of PGS (colors) and CST (x- axis).

However, an antagonistic interaction between the 24-month CST and PGS existed on the multiplicative scale - their joint effect was lower than would be expected given their independent effects (P-value=0.04). There also was a nonsignificant antagonistic interaction on the additive scale (relative excess risk due to interaction:-10.67 (95% CI: (-95.17, 29.18)), Table **3**). Among children with a noncariogenic CST at the 24-month visit (n=70),a high PGS was associated with a 4.83 (1.29,18.2) greater odds of ECC, relative to a low PGS (Table **3**, Figure **2**). Across PGS-strata, the *Streptococcus/Veillonella* CST was always associated with increased ECC odds. Results were similar for the 12-month visit CST (Appendix Table 9). Among cases, the count of decayed or filled primary tooth surfaces by the 60-month visit increased across PGS quartiles, although the trend was not statistically significant (Figure **2**).

**Table 3:**
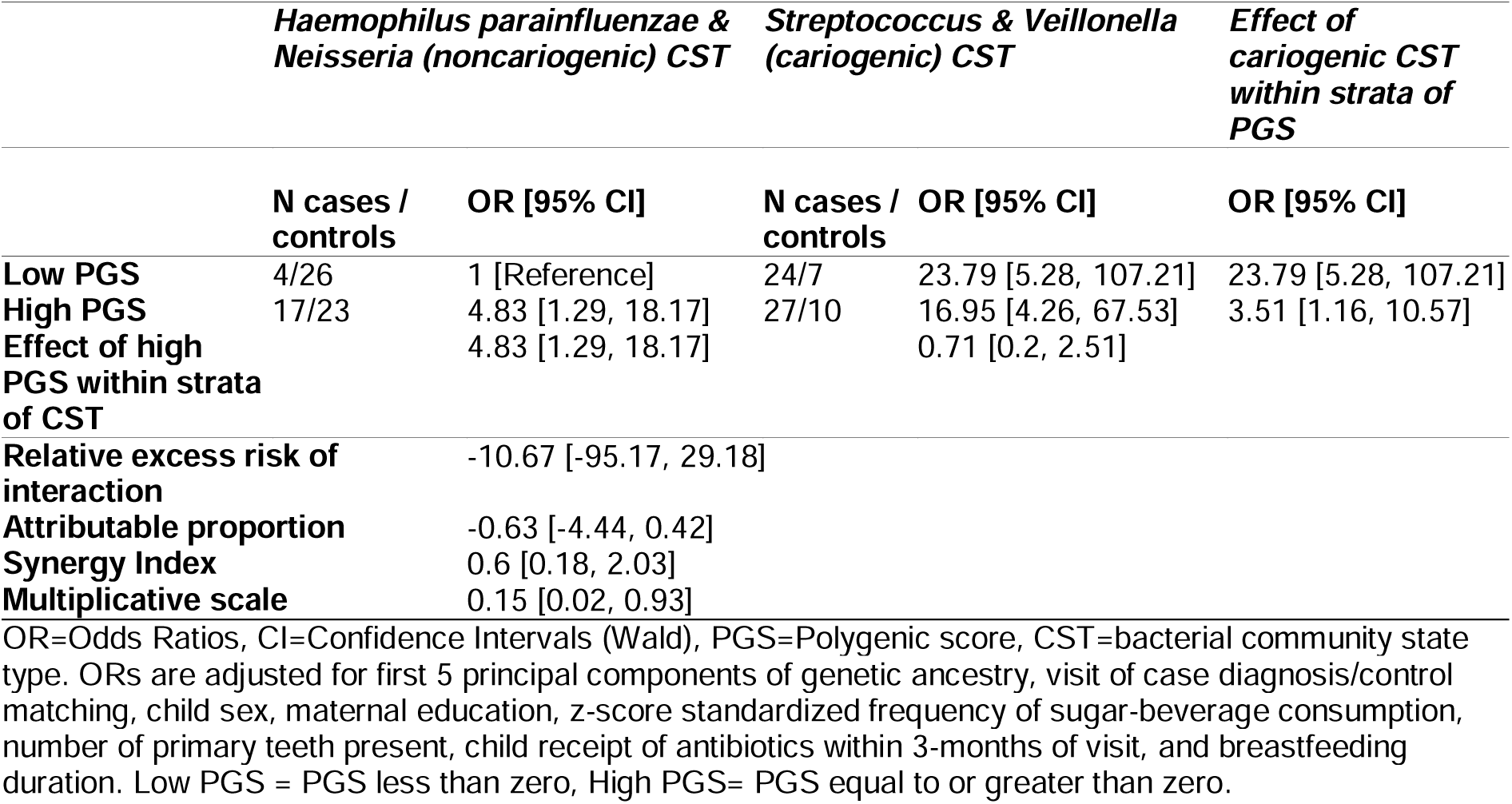
Results from effect modification analysis of dichotomized polygenic score (PGS) and salivary bacterial community state types (CSTs) among 138 early childhood caries (ECC) cases and controls in an incidence-density selected case-control design

|  | *Haemophilus parainfluenzae & Neisseria (noncariogenic) CST* |  | *Streptococcus & Veillonella (cariogenic) CST* |  | *Effect of cariogenic CST within strata of PGS* |
| --- | --- | --- | --- | --- | --- |
|  | N cases / controls | OR [95% CI] | N cases / controls | OR [95% CI] | OR [95% CI] |
| Low PGS | 4/26 | 1 [Reference] | 24/7 | 23.79 [5.28, 107.21] | 23.79 [5.28, 107.21] |
| High PGS | 17/23 | 4.83 [1.29, 18.17] | 27/10 | 16.95 [4.26, 67.53] | 3.51 [1.16, 10.57] |
| Effect of high PGS within strata of CST |  | 4.83 [1.29, 18.17] |  | 0.71 [0.2, 2.51] |  |
| Relative excess risk of interaction |  | -10.67 [-95.17, 29.18] |  |  |  |
| Attributable proportion |  | -0.63 [-4.44, 0.42] |  |  |  |
| Synergy Index |  | 0.6 [0.18, 2.03] |  |  |  |
| Multiplicative scale |  | 0.15 [0.02, 0.93] |  |  |  |

OR=Odds Ratios, CI=Confidence Intervals (Wald), PGS=Polygenic score, CST=bacterial community state type. ORs are adjusted for first 5 principal components of genetic ancestry, visit of case diagnosis/control matching, child sex, maternal education, z-score standardized frequency of sugar-beverage consumption, number of primary teeth present, child receipt of antibiotics within 3-months of visit, and breastfeeding duration. Low PGS = PGS less than zero, High PGS= PGS equal to or greater than zero.

A similar but nonsignificant interaction existed when using salivary *S. mutans* presence instead of CST in models (Appendix Table 10).

## Discussion

Among Appalachian children, we found an antagonistic interaction between a PGS for ECC and the salivary bacterial community. The PGS was significantly associated with ECC only among children with a noncariogenic bacterial community, whereas in children with a cariogenic bacterial community, the PGS was not predictive of ECC. Our work suggests microbial risk factors may modify host genetic impact on ECC.

We found that the PGS was higher in children who experienced ECC, but this association was not statistically significant. Moreover, we observed some heterogeneity in the association by age-at-diagnosis. Our findings fit with a disease model of ECC as multifactorial with a complex polygenic architecture. Caries heritability varies across cohorts, suggesting a polygenic architecture with population heterogeneity (Zeng et al. 2014; Haworth et al. 2018; Shungin et al. 2019). Caries heritability also varies across ages, by longitudinal caries trajectories, between primary and permanent dentition, by tooth type, and by sex (Bretz et al. 2005; Wang et al. 2010; Shaffer, Wang, et al. 2012; Shaffer et al. 2015; Haworth et al. 2020). Given this complex genetic architecture, our analysis is likely underpowered.

Our findings fit with limited previous research on genome-microbiome interactions in ECC. A previous analysis constructed a genetic risk score of three risk SNPs with suggestive *Streptococcus mutans* interactions in a genome-wide scan (Meng et al. 2019). Our PGS did not include the three risk SNPs from this previous analysis but did include SNPs in one of the same genes, galactokinase 2. In the previous analysis, the score was positively associated with ECC only among Appalachian children without *Streptococcus mutans* (Meng et al. 2019). Similarly, in our analysis, the PGS was positively associated with ECC only among those with the noncariogenic CST. Cariogenic microbiomes and host genetics may compete to cause ECC, creating a statistical but not necessarily biological interaction and attenuating genetic associations among those with cariogenic microbiomes (“competing antagonism”, see (VanderWeele and Knol 2011), Supplemental Note). Since environmental exposures can alter measurable genetic associations, genetic analyses should be conducted across populations with varying environmental exposures.

The oral microbiome might mediate as well as modify host genetic effects on ECC. While some work suggests oral microbiome composition is mostly influenced by environmental factors (Shaw et al. 2017), a recent analysis found host genetics explains at least 10% of the variance in the oral microbiome (Liu et al. 2021). Some SNPs in our PGS could affect the microbiome, including SNPs in major histocompatibility complex genes (Andeweg et al. 2021). Taste genes may also affect the microbiome and ECC by changing diets. Our PGS did not contain SNPs in taste genes previously associated with both ECC and the microbiome (i.e. taste 2 receptor member genes) (Cruz de Jesus et al. 2022; Orlova et al. 2023). While we did not observe a significant association between the PGS and the CST, future research in larger cohorts should investigate these mediating pathways.

Our work has several strengths. We had prospective, repeated assessments of both bacterial and dental measures. We used comprehensive summary measures of genetic and bacterial exposures rather than examining single candidate SNPs or bacterial taxa. In the nested case-control analysis, controls were selected from the exact population which gave rise to cases, limiting selection bias from non-representative controls.

Our work also has limitations. We used the largest available GWAS meta-analysis with the same phenotype and patient population to construct our PGS. However, this meta-analysis was relatively small (n<20,000). PGS performance is improved when using weights from larger samples (Lewis and Vassos 2020). We analyzed salivary bacteria, but other microbes may also influence ECC. Loss-to-follow-up also occurred in COHRA2. We performed an inverse probability weighting sensitivity analysis, but selection bias could still exist. Given the population heterogeneity of ECC and the small sample size, our analysis is underpowered, and future replication efforts in larger samples or meta-analyses are necessary.

Because the GWAS meta-analysis we used for SNP weights was conducted among European ancestry children, we limited our analysis to children with a maternal-reported race of white. While race and genetic ancestry are not synonymous (Khan et al. 2022), maternal report of child race and genetic ancestry were highly concordant in COHRA2. Our findings may not generalize to other genetic ancestries, especially since microbiomes also vary across human populations. Over-representation of European-ancestry individuals in GWAS and subsequent PGS applications could exacerbate health disparities in ECC (Martin et al. 2019). Future analyses of genome-microbiome interactions in ECC should be conducted in diverse populations.

Our work suggests strong environmental risk factors, such as the oral microbiome, can modify the measurable effect of host genetic risk. However, as the bacterial community was associated with ECC across genetic risk strata, interventions targeting cariogenic microbiomes in early childhood can be broadly applied across genetic risk strata.

## Data Availability

The 16S rRNA gene amplicon sequencing data from the COHRA2 study is publicly available at the PRJNA752888 repository. Phenotype and host genomic data for the COHRA2 study are available at dbGaP phs001591.v1.p1 upon application.

https://www.ncbi.nlm.nih.gov/bioproject/PRJNA752888

## Acknowledgments

This study was funded by the National Institutes for Health National Institute for Dental and Craniofacial Research. The COHRA2 cohort, including data collection and microbial sequencing was funded through grant R01 DE014899. The genotyping of the COHRA2 cohort was supported through grant. The work of FB was supported through grant F31 DE029992.

## Author contributions

Freida Blostein contributed to the conception, analysis and interpretation of this paper and drafted the manuscript.

Tianyu Zou contributed to the analysis and interpretation and critically revised the manuscript.

Deesha Bhaumik contributed to the analysis and critically revised the manuscript.

Elizabeth Salzman contributed to the acquisition of the data and critically revised the manuscript.

Kelly M. Bakulski contributed to the analysis and interpretation and critically revised the manuscript.

John R. Shaffer contributed to the conception, design, and interpretation as well as critically revising the manuscript.

Mary L. Marazita contributed to the conception, design and interpretation as well as critically revising the manuscript.

Betsy Foxman contributed to the conception, design and interpretation as well as critically revising the manuscript.

All authors gave their final approval and agree to be accountable for all aspects of the work.

## Data availability

The 16S rRNA amplicon sequencing data used in this analysis is available at PRJNA752888. Genotype and phenotype data for COHRA2 is available from dbGAP upon reasonable request (phs001591.v1.p1). The base data statistics used in the creation of the polygenic score are available at doi:10.6084/m9.figshare.21890670.

## References

Alli BY. 2021. InteractionR: An R package for full reporting of effect modification and interaction. Software Impacts. 10:100147.

Andeweg SP, Keşmir C, Dutilh BE. 2021. Quantifying the Impact of Human Leukocyte Antigen on the Human Gut Microbiota. mSphere. 6(4):e0047621.

Bezamat M, Souza JF, Silva FMF, Corrêa EG, Fatturi AL, Brancher JA, Carvalho FM, Cavallari T, Bertolazo L, Machado-Souza C, et al. 2021. Gene-environment interaction in molar-incisor hypomineralization. PLOS ONE. 16(1):e0241898.

Blostein F, Bhaumik D, Davis E, Salzman E, Shedden K, Duhaime M, Bakulski KM, McNeil DW, Marazita ML, Foxman B. 2022. Evaluating the ecological hypothesis: early life salivary microbiome assembly predicts dental caries in a longitudinal case-control study. Microbiome. 10(1):240. doi:10.1186/s40168-022-01442-5.

Boraas JC, Messer LB, Till MJ. 1988. A Genetic Contribution to Dental Caries, Occlusion, and Morphology as Demonstrated by Twins Reared Apart. J Dent Res. 67(9):1150–1155.

Bretz WA, Corby PM, Schork NJ, Robinson MT, Coelho M, Costa S, Melo Filho MR, Weyant RJ, Hart TC. 2005. Longitudinal Analysis of Heritability for Dental Caries Traits. J Dent Res. 84(11):1047–1051.

Cruz de Jesus V, Mittermuller BA, Hu P, Schroth RJ, Chelikani P. 2022. Genetic variants in taste genes play a role in oral microbial composition and severe early childhood caries. iScience. 25(12):105489.

Das S, Forer L, Schönherr S, Sidore C, Locke AE, Kwong A, Vrieze SI, Chew EY, Levy S, McGue M, et al. 2016. Next-generation genotype imputation service and methods. Nat Genet. 48(10):1284–1287.

Fleming E, Afful J. 2018. Prevalence of total and untreated dental caries among youth: United states, 2015-2016. NCHS Data Brief.(307):1–8.

Grier A, Myers JA, O’Connor TG, Quivey RG, Gill SR, Kopycka-Kedzierawski DT. 2020. Oral Microbiota Composition Predicts Early Childhood Caries Onset. J Dent Res. 100(6):599–607.

Haworth S, Esberg A, Lif Holgerson P, Kuja-Halkola R, Timpson NJ, Magnusson PKE, Franks PW, Johansson I. 2020. Heritability of Caries Scores, Trajectories, and Disease Subtypes. J Dent Res. 99(3):264–270.

Haworth S, Shungin D, van der Tas JT, Vucic S, Medina-Gomez C, Yakimov V, Feenstra B, Shaffer JR, Lee MK, Standl M, et al. 2018. Consortium-based genome-wide meta-analysis for childhood dental caries traits. Hum Mol Genet. 27(17):3113–3127.

Holmes I, Harris K, Quince C. 2012. Dirichlet Multinomial Mixtures: Generative Models for Microbial Metagenomics. PLoS ONE. 7(2):e30126.

Khan AT, Gogarten SM, McHugh CP, Stilp AM, Sofer T, Bowers ML, Wong Q, Cupples LA, Hidalgo B, Johnson AD, et al. 2022. Recommendations on the use and reporting of race, ethnicity, and ancestry in genetic research: Experiences from the NHLBI TOPMed program. Cell Genomics. 2(8):100155.

Knol MJ, VanderWeele TJ. 2012. Recommendations for presenting analyses of effect modification and interaction. Int J Epidem. 41(2):514–520.

Lee SH, Goddard ME, Wray NR, Visscher PM. 2012. A Better Coefficient of Determination for Genetic Profile Analysis. Genet Epidemiol. 36(3):214–224.

Lewis CM, Vassos E. 2020. Polygenic risk scores: from research tools to clinical instruments. Genome Med. 12(1):44.

Liu X, Tong X, Zhu J, Tian L, Jie Z, Zou Y, Lin X, Liang H, Li W, Ju Y, et al. 2021. Metagenome-genome-wide association studies reveal human genetic impact on the oral microbiome. Cell Discovery. 7(1):117.

Martin AR, Kanai M, Kamatani Y, Okada Y, Neale BM, Daly MJ. 2019. Clinical use of current polygenic risk scores may exacerbate health disparities. Nat Genet. 51(4):584–591.

Martins MT, Sardenberg F, Bendo CB, Abreu MH, Vale MP, Paiva SM, Pordeus IA. 2017. Dental caries remains as the main oral condition with the greatest impact on children’s quality of life. PLOS ONE. 12(10):e0185365.

Mayhew AJ, Meyre D. 2017. Assessing the Heritability of Complex Traits in Humans: Methodological Challenges and Opportunities. Curr Genomics. 18(4):332–340.

Meng Y, Wu T, Billings R, Kopycka-Kedzierawski DT, Xiao J. 2019. Human genes influence the interaction between Streptococcus mutans and host caries susceptibility: a genome-wide association study in children with primary dentition. Int J Oral Sci. 11(2):19.

Neiswanger K, McNeil DW, Foxman B, Govil M, Cooper ME, Weyant RJ, Shaffer JR, Crout RJ, Simhan HN, Beach SR, et al. 2015. Oral Health in a Sample of Pregnant Women from Northern Appalachia (2011-2015). Int J of Dentistry. 2015:1–12.

Orlova E, Dudding T, Chernus JM, Alotaibi RN, Haworth S, Crout RJ, Lee MK, Mukhopadhyay N, Feingold E, Levy SM, et al. 2023. Association of early childhood caries with bitter taste receptors: A meta-analysis of genome-wide association studies and transcriptome-wide association study. Genes. 14(1):59.

Pitts NB, Zero DT, Marsh PD, Ekstrand K, Weintraub JA, Ramos-Gomez F, Tagami J, Twetman S, Tsakos G, Ismail A. 2017. Dental caries. Nat Rev Disease Primers. 3(1):17030.

Shaffer JR, Feingold E, Wang X, TCuenco K, Weeks DE, DeSensi RS, Polk DE, Wendell S, Weyant RJ, Crout R, et al. 2012. Heritable patterns of tooth decay in the permanent dentition: principal components and factor analyses. BMC Oral Health. 12(1):7.

Shaffer JR, Wang X, DeSensi RS, Wendell S, Weyant RJ, Cuenco KT, Crout R, McNeil DW, Marazita ML. 2012. Genetic Susceptibility to Dental Caries on Pit and Fissure and Smooth Surfaces. Caries Res. 46(1):38–46.

Shaffer JR, Wang X, McNeil DW, Weyant RJ, Crout R, Marazita ML. 2015. Genetic Susceptibility to Dental Caries Differs between the Sexes: A Family-Based Study. Caries Res. 49(2):133–140.

Shaw L, Ribeiro ALR, Levine AP, Pontikos N, Balloux F, Segal AW, Roberts AP, Smith AM. 2017. The Human Salivary Microbiome Is Shaped by Shared Environment Rather than Genetics: Evidence from a Large Family of Closely Related Individuals. mBio. 8(5):e01237–17.

Shungin D, Haworth S, Divaris K, Agler CS, Kamatani Y, Keun Lee M, Grinde K, Hindy G, Alaraudanjoki V, Pesonen P, et al. 2019. Genome-wide analysis of dental caries and periodontitis combining clinical and self-reported data. Nat Commun. 10(1):2773.

Slayton RL, Cooper ME, Marazita ML. 2005. Tuftelin, Mutans Streptococci, and Dental Caries Susceptibility. J Dent Res. 84(8):711–714.

Takahashi N, Nyvad B. 2010. The Role of Bacteria in the Caries Process. J Dent Res. 90(3):294–303.

the Haplotype Reference Consortium. 2016. A reference panel of 64,976 haplotypes for genotype imputation. Nat Genet. 48(10):1279–1283.

VanderWeele TJ, Knol MJ. 2011. Remarks on Antagonism. Am J Epidemiol. 173(10):1140–1147.

Wand H, Lambert SA, Tamburro C, Iacocca MA, O’Sullivan JW, Sillari C, Kullo IJ, Rowley R, Dron JS, Brockman D, et al. 2021. Improving reporting standards for polygenic scores in risk prediction studies. Nature. 591(7849):211–219.

Wang X, Shaffer JR, Weyant RJ, Cuenco KT, DeSensi RS, Crout R, McNeil DW, Marazita ML. 2010. Genes and Their Effects on Dental Caries May Differ between Primary and Permanent Dentitions. Caries Res. 44(3):201–208.

Wang X, Willing MC, Marazita ML, Wendell S, Warren JJ, Broffitt B, Smith B, Busch T, Lidral AC, Levy SM. 2012. Genetic and Environmental Factors Associated with Dental Caries in Children: The Iowa Fluoride Study. Caries Res. 46(3):177–184.

Zeng Z, Feingold E, Wang X, Weeks DE, Lee M, Cuenco KT, Broffitt B, Weyant RJ, Crout R, McNeil DW, et al. 2014. Genome-Wide Association Study of Primary Dentition Pit-and-Fissure and Smooth Surface Caries. Caries Res. 48(4):330–338.

Zou G. 2004. A Modified Poisson Regression Approach to Prospective Studies with Binary Data. Am J Epidemiol. 159(7):702–706.

Zou GY. 2008. On the Estimation of Additive Interaction by Use of the Four-by-two Table and Beyond. Am J Epidemiol. 168(2):212–224.

