## Supplemental Materials for "Bacterial community modifies host genetics effect on early childhood caries"

Bacterial community modifies host genetics effect on early childhood caries: Supplemental Methods and Results + Appendix Figures and Tables

**Authors:**

Freida Blostein${}^{1}$, Tianyu Zou${}^{23}$, Deesha Bhaumik${}^{1}$, Elizabeth Salzman${}^{1}$, Kelly M. Bakulski^1^, John R. Shaffer${}^{23}$, Mary L. Marazita${}^{234}$, Betsy Foxman${}^{1}$

**Affiliations:**

${}^{1}$ Department of Epidemiology, University of Michigan School of Public Health, University of Michigan, Ann Arbor, Michigan, United States of America

${}^{2}$ Department of Oral and Craniofacial Sciences, Center for Craniofacial and Dental Genetics, School of Dental Medicine, University of Pittsburgh, Pittsburgh, Pennsylvania, United States of America

${}^{3}$ Department of Human Genetics, School of Public Health, University of Pittsburgh, Pittsburgh, Pennsylvania, United States of America

${}^{4}$ Clinical and Translational Sciences Institute, and Department of Psychiatry, School of Medicine, University of Pittsburgh, Pittsburgh, Pennsylvania, United States of America

### Supplemental Methods

#### COHRA2 cohort eligibility

Healthy women $\geq$ 18 years of age, who were fluent in English and in the 12th to 29th week of a singleton pregnancy were eligible for inclusion. Women who thought they might leave West Virginia or southwestern Pennsylvania soon and those without a reliable telephone contact were ineligible

#### In-depth description of human genetic data bioinformatics

For SNP weights, we used the effect estimates from a previous, independent genome-wide association study meta-analysis of 22 European-ancestry cohorts (n=17,666 children aged 2.50- to 11.99-years) with a phenotype of any decay in primary dentition (base data set) [1]. In this meta-analysis, 12,866,520 SNPs were tested, with the lowest p value being 4.13E-8 [1]. To promote harmonization with the COHRA2 data (target data set), we removed SNPs with a minor allele frequency $\leq$ 0.01 and ambiguous SNPs. No duplicate SNPs existed in the COHRA2 data and no individuals were related. After quality-control, 6,044,259 SNPs in the target data overlapped with the base data. For clumping, the value 0.1 was used for the squared correlation threshold, and the window size of clumping was set to 250 KB. PGS were calculated at 7 $P$-value thresholds: 0.001, 0.05, 0.1, 0.2, 0.3, 0.4 and 0.5. To improve interpretability, PGSs were z-score standardized. To facilitate epidemiological interpretations, we dichotomized the z-score standardized PGS into high PGS ($\geq0$) and low PGS ($<0$) for some statistical analyses.

#### In-depth description of incidence-density sampling

Appendix Figure 1 shows an illustrative random sample of follow-up time in the COHRA2 cohort. Children are shown as lines, with follow-up time represented by the length of the line across the x-axis. Person-time for children at-risk of ECC is shown in either black (for children never diagnosed with ECC) or blue (for children ever diagnosed with ECC). After a child is diagnosed with ECC, they are no longer at-risk for ECC (age at diagnosis shown as orange Xs, time not at-risk shown in orange). In incidence density sampling, children diagnosed at the 36-month visit (for example, orange Xs within grey rectangle in Appendix Figure 1B) are selected into the nested case-control subset. All children still at-risk of first diagnosis of primary tooth decay (black and blue lines within grey rectangle) are eligible to be selected as risk-set controls, while children already diagnosed with primary tooth decay (orange lines within grey rectangle) are not eligible.

In an incidence-density sampled design, individuals can be selected as both cases and controls or as controls in multiple risk-sets. In our sampling, one individual was selected as both a case and a control and one individual was selected as a control in two risk-sets. For statistical analyses, these two children are therefore represented twice each in the data. Thus, the complete number of cases and controls in the analysis is 148, but these cases and controls represent a total of unique individuals 146. Similarly, though there were 138 cases and controls with data available at 24-months, these are composed of 136 unique individuals, and for 12-months 134 cases and controls represent 132 unique individuals.

Importantly, sampling for this incidence density sampled case-control subset was conducted before all the follow-up data for the cohort had been completed, therefore not all cases of early childhood caries from the cohort are represented in the nested subset. Specifically, the cases diagnosed at earlier ages are better represented in the nested subset than cases diagnosed at later ages (Appendix Table 2).

In an incidence-density sampled case-control design, the OR estimates the IRR. In any case-control design with matching, it is important to control for matching variables to prevent confounding. When matched sets are one-to-one, conditional regression is often used. However, if matched sets are not one-to-one, it can be more statistically efficient to us unconditional matching while controlling for matching variables. In this analysis, cases were matched to controls on visit of case diagnosis but there were multiple cases and controls within these risk-sets, thus we used unconditional regression while controlling for visit of matching [2].

In our analyses of incidence rates of ECC, all person-time up to first diagnosis of primary tooth decay is compared to person time of undiagnosed children (blue compared to black lines). In our analyses of prevalence of ECC, all children with prevalent ECC are compared to children without prevalent dental decay (orange compared to black and blue lines).

#### Processing of saliva and bioinformatics for microbial genetic data analyses

Bacterial DNA was extracted from aliquots of saliva. Library preparation and sequencing of the 16S rRNA V4 amplicon was performed by the Michigan Microbial Systems Molecular Biology Laboratory using previously validated protocols [3]. DNA extraction was performed using the Eppedorf EpMotion liquid handling system following the Qiagen MagAttract PowerMicrobiome kit protocol. The V4 variable region was amplified from extracted DNA using barcoded dual-index primers and sequenced on the Illumina MiSeq platform using the MiSeq Reagent Kit V2 500 cycles. Each plate of samples was submitted with a positive mock community control, a DNA extraction kit control, and a negative water control. Reads were processed to amplicon sequence variants (ASVs) using DADA2 (version 1.14.1) [4] and the Human Oral Microbiome Database (HOMD) version 15.2 [5]. After examining quality plots for forward and reverse reads, reads were trimmed at the 240th and 200th nucleotide position respectively, or at the first instance of a quality score <= to 11. The following parameters were used for filtering reads: a maximum of 2 expected errors for both forward and reverse reads and no ambiguous bases. Errors were learned (independently for each run) using 1 million bases. The forward and reverse reads were denoised and then forward and reverse reads were merged together. To identify contaminants, we used the R package decontam (version 1.8.0) [6]. We excluded 6 samples with less than 1,000 reads. Further details are available in the original publication describing the incident-density sample subsample (Blostein 2022, doi:[10.1186/s40168-022-01442-5](https://doi.org/10.1186/s40168-022-01442-5)). We filtered out ASVs that were both 1) present in less than 5% of the samples and 2) represented less than 5% of the relative abundance in samples in which they were present. This left a total of 273 ASVs used in creating community state types (CSTs).

To determine the number of community state types (CSTs), we fit ten Dirichlet multinomial models, varying the number of Dirichlet components (i.e., CSTs) from 1 to 10. We calculated the Laplace measure of fit for each model and plotted against k, identifying k = 6 as the best model.

Alpha diversity indexes were calculated using the vegan package version 2.5.7. The Chao1 index estimates species richness, or number of microbial populations observed in each sample. The Shannon index combines information on richness and evenness of microbial populations.

#### Additional description of variable creation

Prenatal maternal illness was derived from the question “When were you last sick with a flue-like illness, a really bad cold, fever, a rash or muscle or joint aches?” and was dichotomized into within 1-month of prenatal visit versus over 1-month prior to prenatal visit. Prenatal maternal antibiotic use was based on the question ‘During this pregnancy, have you taken any antibiotics?’ with the dichotomous response yes versus no. Prenatal maternal smoking behavior was based on the question “Thinking back to the three months before you were pregnant up to now, have you smoked cigarettes?” with the dichotomous response yes versus no.

At visits and between-visit phone call interviews, mothers were asked if their child had received any antibiotics, and how long ago the antibiotic had been stopped. Based on the length of time since antibiotic course had been stopped and the date of the visit and call, we calculated a binary variable for any antibiotics received within 3-months of each visit. At in-person visits and during phone interviews, mothers were also administered a food frequency questionnaire. This questionnaire assessed the frequency with which children consumed sugar-sweetened beverages including flavored waters, sports drinks, sodas, powder-mix drinks, coffee, tea, energy drinks and meal replacement drinks. Available codes and responses were ‘1 - Never or once’, ‘2 - every few days’, ‘3 - once a day’, or ‘4 - several times a day’. For each of the above drinks, mothers were asked if the beverage was typically sugar-sweetened, artificially-sweetened (diet), or unsweetened; we only considered responses where the beverage was sugar-sweetened. Mothers were also asked about the frequency of child juice and watered-down juice consumption. We created a single summed score for sugar-sweetened-beverage and juice consumption by summing the reported frequency codes of these beverages and z-score standardizing. At in-person visits and phone call interviews, mothers were asked if they had ever breastfed and if they were currently breastfeeding. If they reported ever breastfeeding but no current breastfeeding, they were asked the child’s age in months and weeks of breastfeeding cessation. Some mothers did not report the child’s age at which they stopped breastfeeding, for these children the age of breastfeeding cessation was calculated as the midpoint between the age of last reported current breastfeeding and the age at first report of no current breastfeeding. Using the age of breastfeeding cessation, we created a categorized duration of breastfeeding variable (never breastfed, 6-months or less, >6 months & <= 12 months, >12 months and <= 24 months, and >24 months). We also created for each visit a dichotomous current breastfed variable (yes versus no).

#### Inverse probability weighting sensitivity model

We calculated inverse probability of treatment and censoring weights, $W^{AC}=W^{A}*W^{C}$. $W^{A}$ is defined as $W^{A}=1/f\left( A|L \right)$ such that $f\left( A|L \right)=Pr\left[ HighPGS|L \right]$ for those individuals with a high PGS, and $f\left( A|L \right)=Pr\left[ LowPGS|L \right]$ for those individuals with a low PGS. $W^{C}$ is defined as $W^{C}=1/Pr\left[ Uncensored|PGS,L \right]$. For both treatment and outcome models, $L$ was defined as the full health behaviors model described above, plus the additional variables of maternal age at prenatal visit and an indicator for if the focal child was the mother’s firstborn child, as these were strongly associated with the probability of censorship, and it is most efficient to keep all variables in both the treatment and censoring model. These variables were ascertained from maternal interview at the prenatal visit. $W^{AC}$ were then used as weights in marginal models for the incidence rate and prevalence rates, and the robust variance estimate was used.

Since the treatment as defined (high vs low PGS) can only take on two values, stabilized weights and non-stabilized weights will result in the same estimate. For the incidence rate Poisson model, children who were lost-to-follow-up before completing any dental exams with emerged primary teeth were considered censored.

#### R packages used in the creation and visualization of tables and figures

The following R packages were used in this analysis for data cleaning and analysis: broom, tidyverse, dplyr, stringr, sjlabelled, MASS, vegan, DirichletMultinomial [7–14]. The following R packages were used for the creation and formating of tables and figures: gtsummary, huxtable, flextable, ggplot2, ggpubr [15–19].

### Supplemental Results

#### Inverse probability weighting sensitivity model

After applying inverse probability of treatment weights, the incidence rate ratio estimate from the marginal model was slightly attenuated from that estimated in the conditional models controlling for known covariates (IRR: 1.06 95% CI: (0.8, 1.41)). Further application of censoring weights resulted in a similar estimate, suggesting a small effect of selection bias (IRR: 1.05 95% CI: 0.8, 1.4).

For the cross-sectional prevalence ratio (PR) Poisson models, children who were lost-to-follow-up before the respective visit (3- or 5-year visit) were considered censored. Applying inverse-probability of exposure weights in marginal models reduced the effect estimates from those estimated in the conditional model (3-year PR: 1.49 95%CI 0.94, 2.36, 5-year PR 1.23 95% CI: 0.87, 1.74). Additionally applying censoring weights slightly reduced effect estimates (3-year PR: 1.38 95%CI 0.86, 2.22, 5-year PR 1.22 95% CI: 0.87, 1.72).

### Appendix Figures

#### Appendix Figure 1: Study design A) Distribution of all available visits (light grey), visits with a child’s first diagnosis of ECC in entire cohort (dark grey), and visits with first diagnosis of ECC in nested case-control subset (black by age of child at visit. Black lines denote the intended age of each visit in cohort procedures. B) Illustrative random sample of individual children’s available follow-up and potential selection into nested case-control subset. Each child is represented as a line extending from birth to their age at their last available visit. Children who were never diagnosed with ECC are denoted in black. Orange Xs denote the age at first diagnosis of ECC, time at-risk of diagnosis with first ECC shown in blue, time after first diagnosis of ECC shown in orange. In incidence density sampling, children diagnosed at the 36-month visit (for example, orange Xs within grey rectangle) are selected into the nested case-control subset. All children still at-risk of first diagnosis of ECC (black and blue lines within grey rectangle) are eligible to be selected as risk-set controls, children already diagnosed with ECC (orange lines within grey rectangle) are not eligible. In our analyses of incidence rates of first ECC, all person-time up to first diagnosis of ECC and all person time of undiagnosed children is considered (blue and black lines). In our analyses of prevalence of ECC, all children with prevalent dental decay are compared to children without prevalent dental decay (orange compared to black and blue lines).

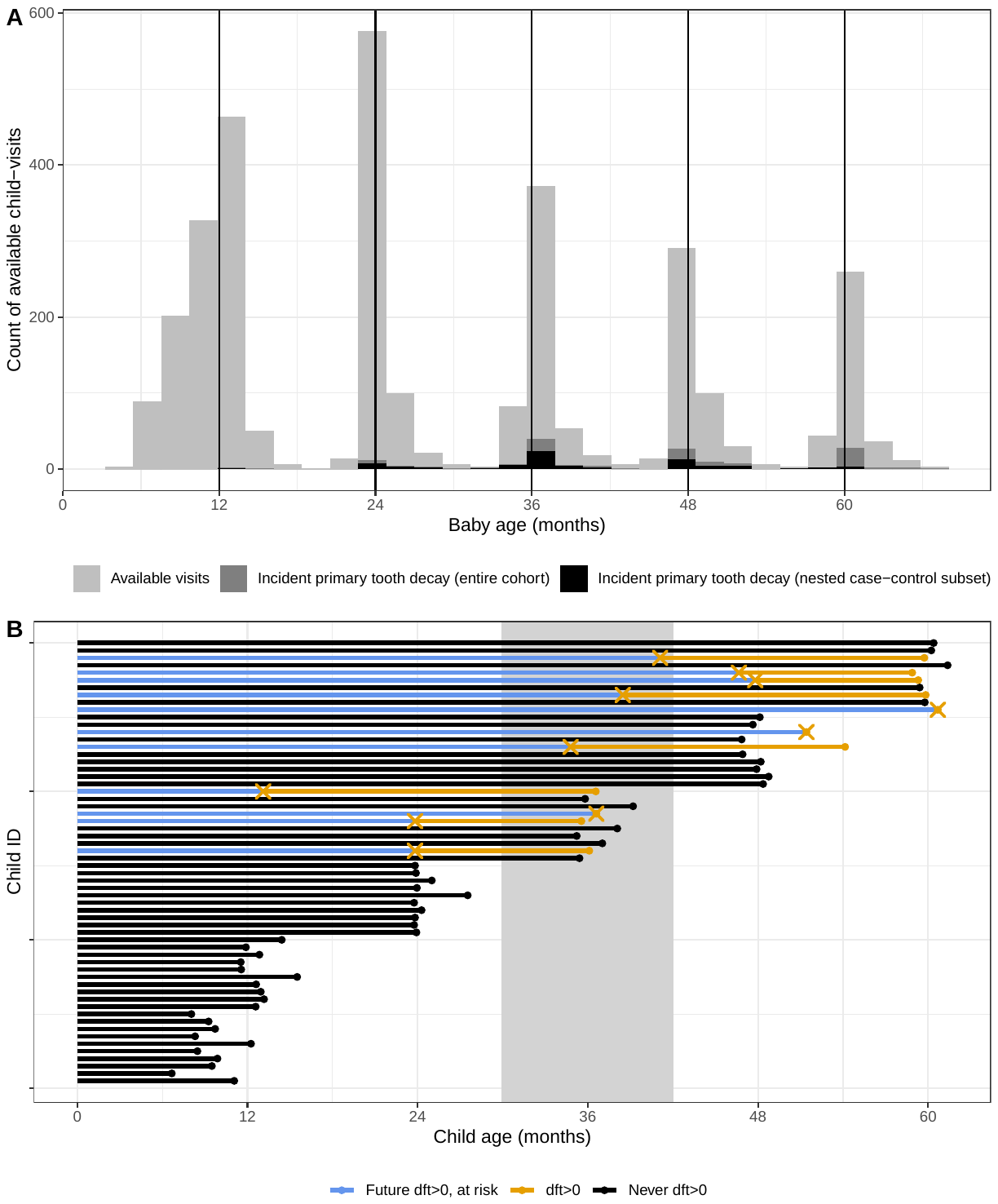

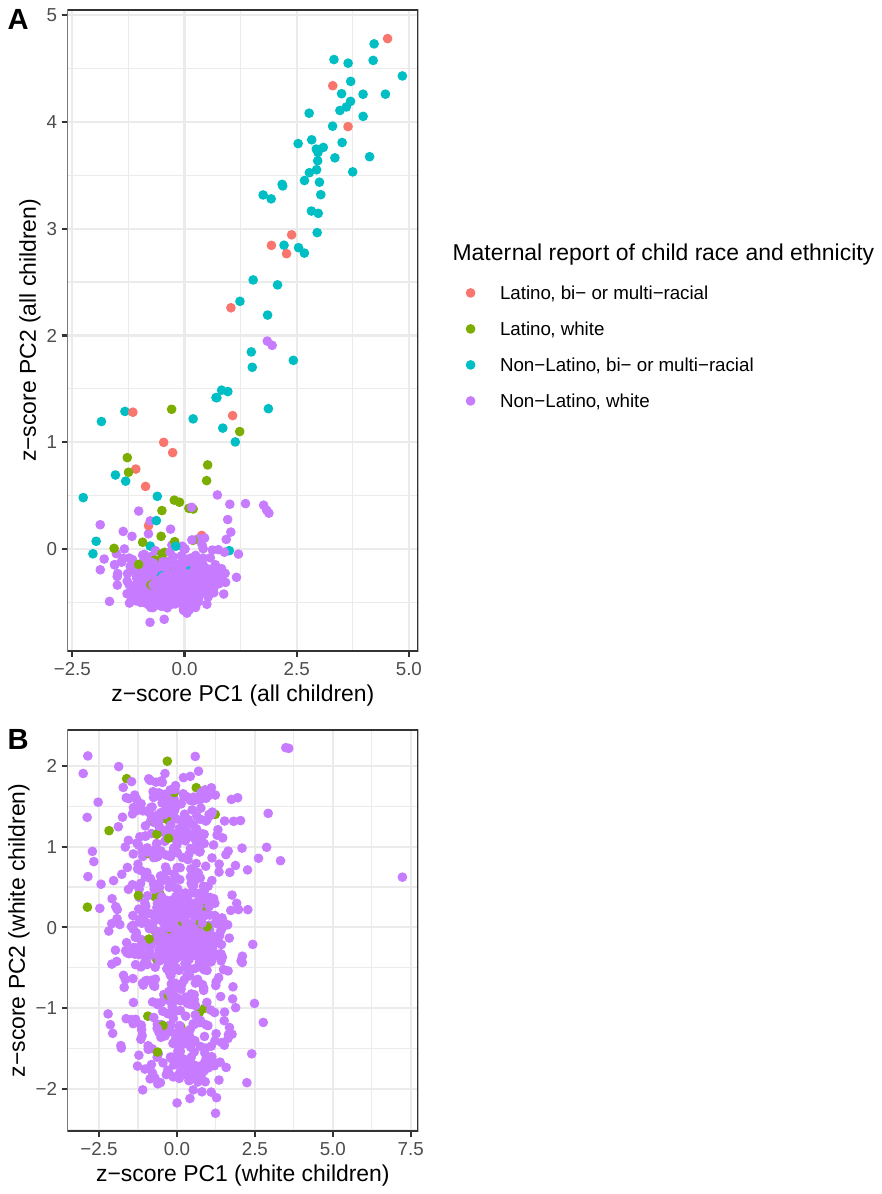

#### Appendix Figure 2: Correspondence of principal components of genetic ancestry with maternal report of child race among Appalachian children in the Center for Oral Health in Appalachia 2 longitudinal cohort study

#### Appendix Figure 3: Exclusion of participants from the analytic subset among the COHRA2 longitudinal cohort and selection into the case-control nested subset

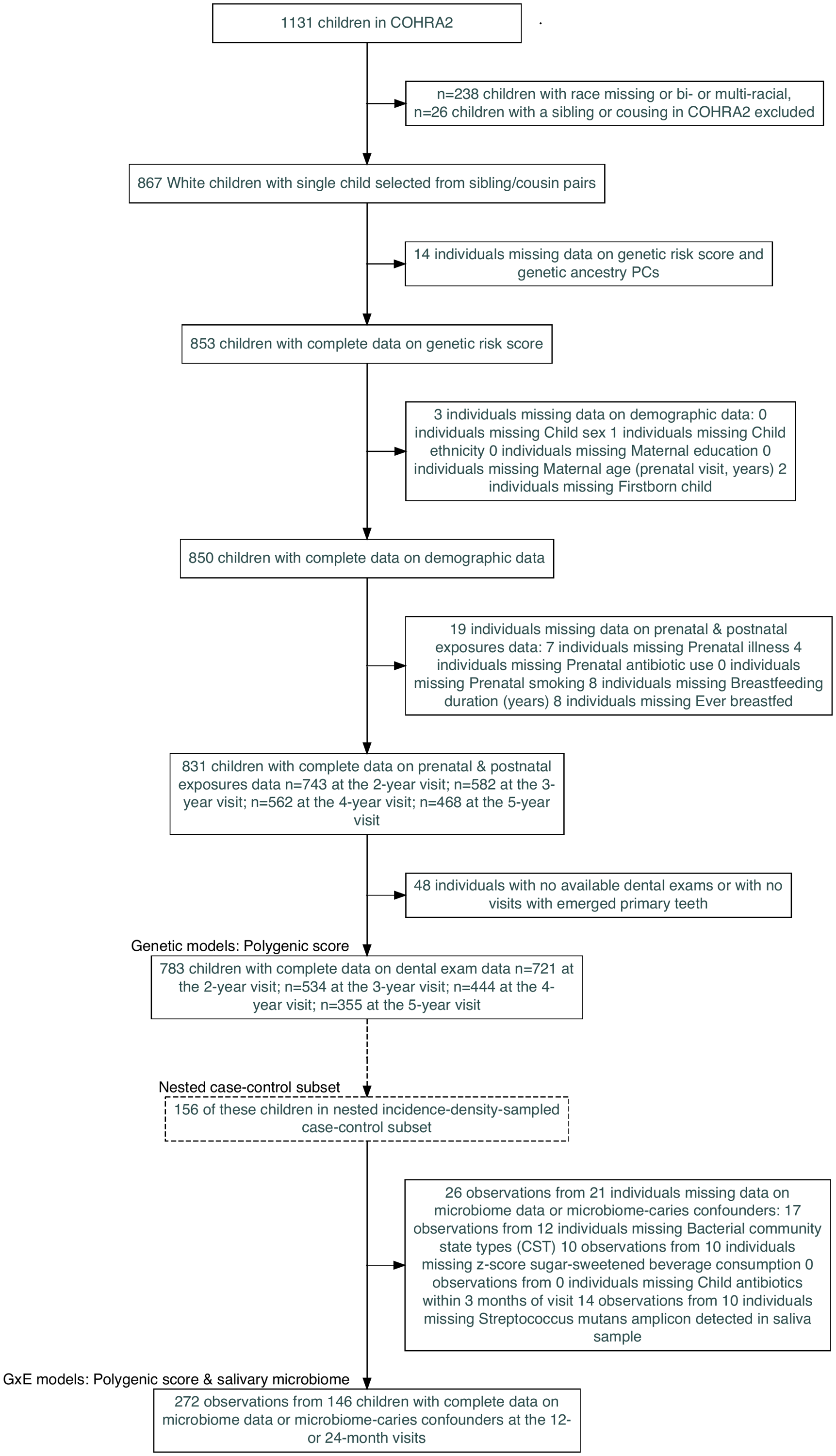

#### Appendix Figure 4: $\boldsymbol{R}^{\boldsymbol{2}}$ values for prevalent ECC and PGS created using various *P*-value thresholds at the 36-, 48- and 60-month visits among 783 Appalachian children in the Center for Oral Health in Appalachia 2 longitudinal cohort study

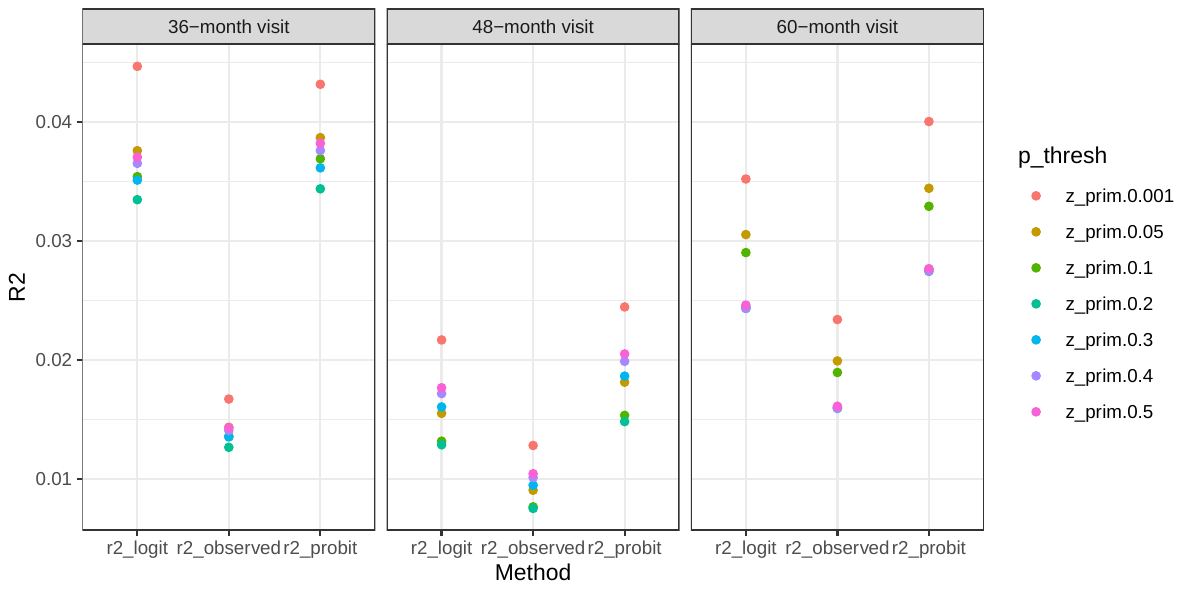

#### Appendix Figure 5: Distribution of PGS for ECC created using various *P*-value thresholds among 783 Appalachian children in the Center for Oral Health in Appalachia 2 longitudinal cohort study

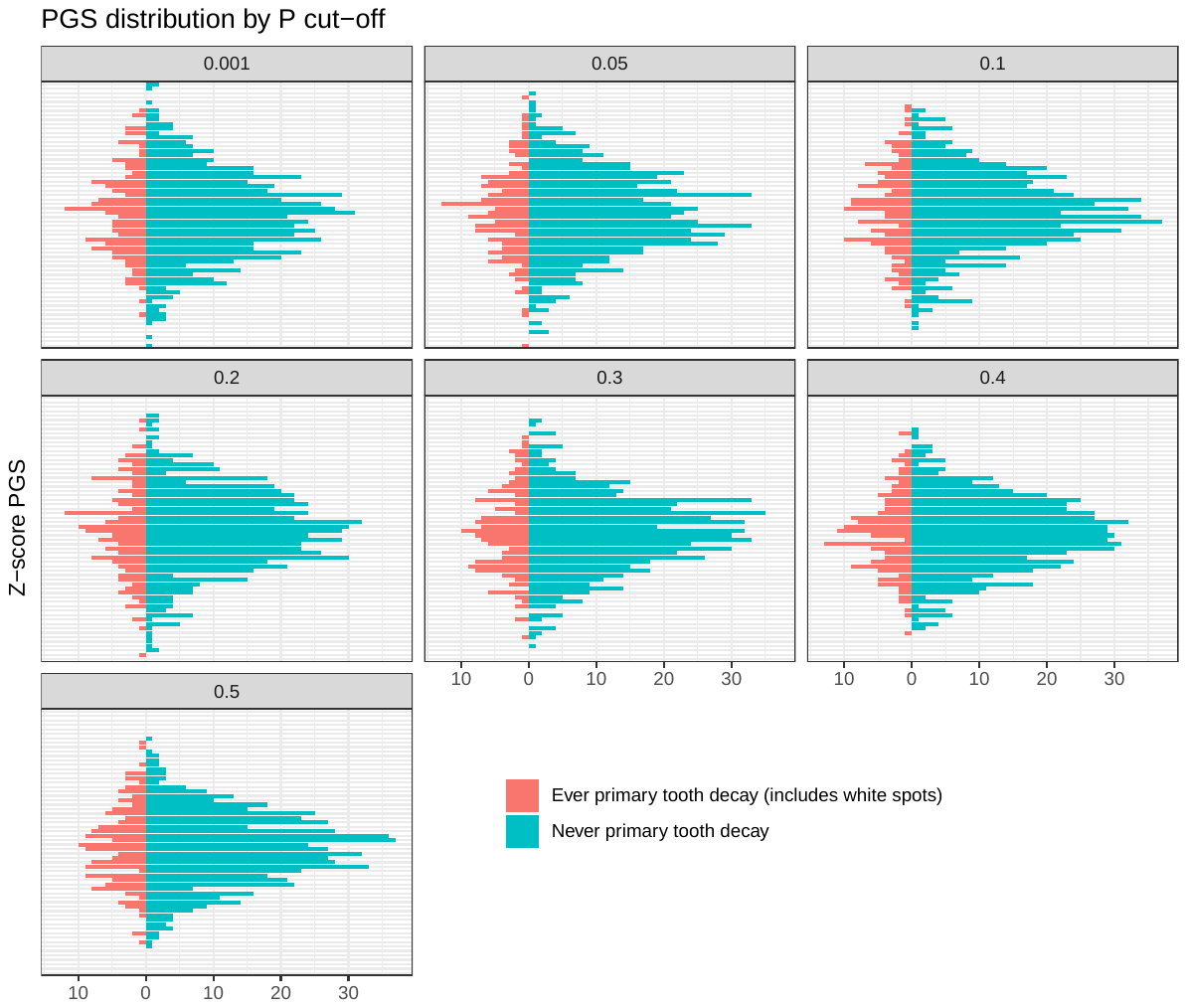

#### Appendix Figure 6: Association between PGS for ECC and age-at-diagnosis of ECC using ECC excluding white spots as definition of ECC among 783 Appalachian children in the Center for Oral Health in Appalachia 2 longitudinal cohort study

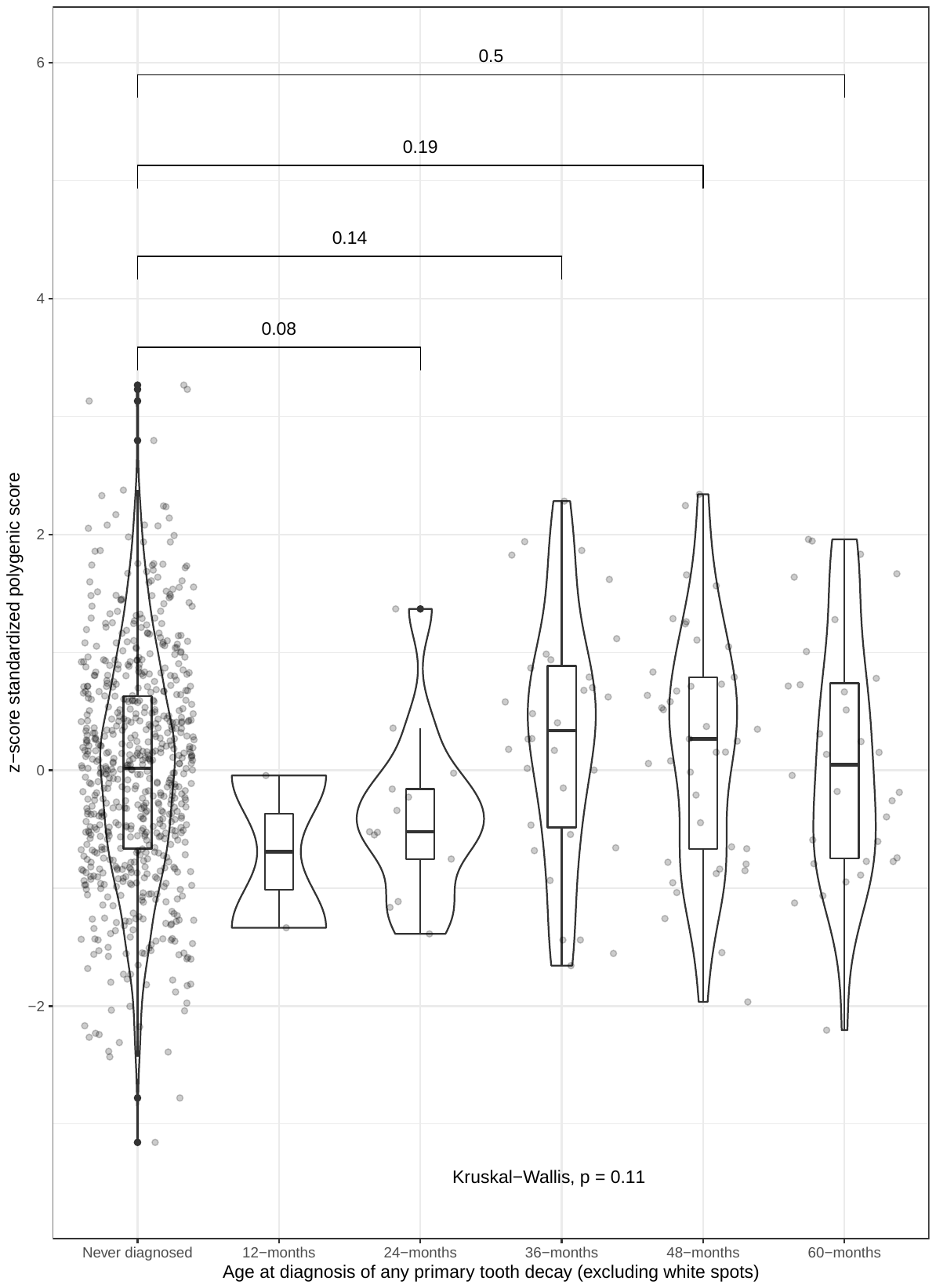

#### Appendix Figure 7: A) Distribution of z-score standardized sugar-sweetened beverage consumption by salivary microbiome community state type and visit B) Distribution of z-score standardized PGS for ECC by salivary microbiome community state type and visit C) Distribution of z-score standardized PGS for ECC by *Streptococcus mutans* amplicon detection and visit among 148 Appalachian children in the Center for Oral Health in Appalachia 2 longitudinal cohort study

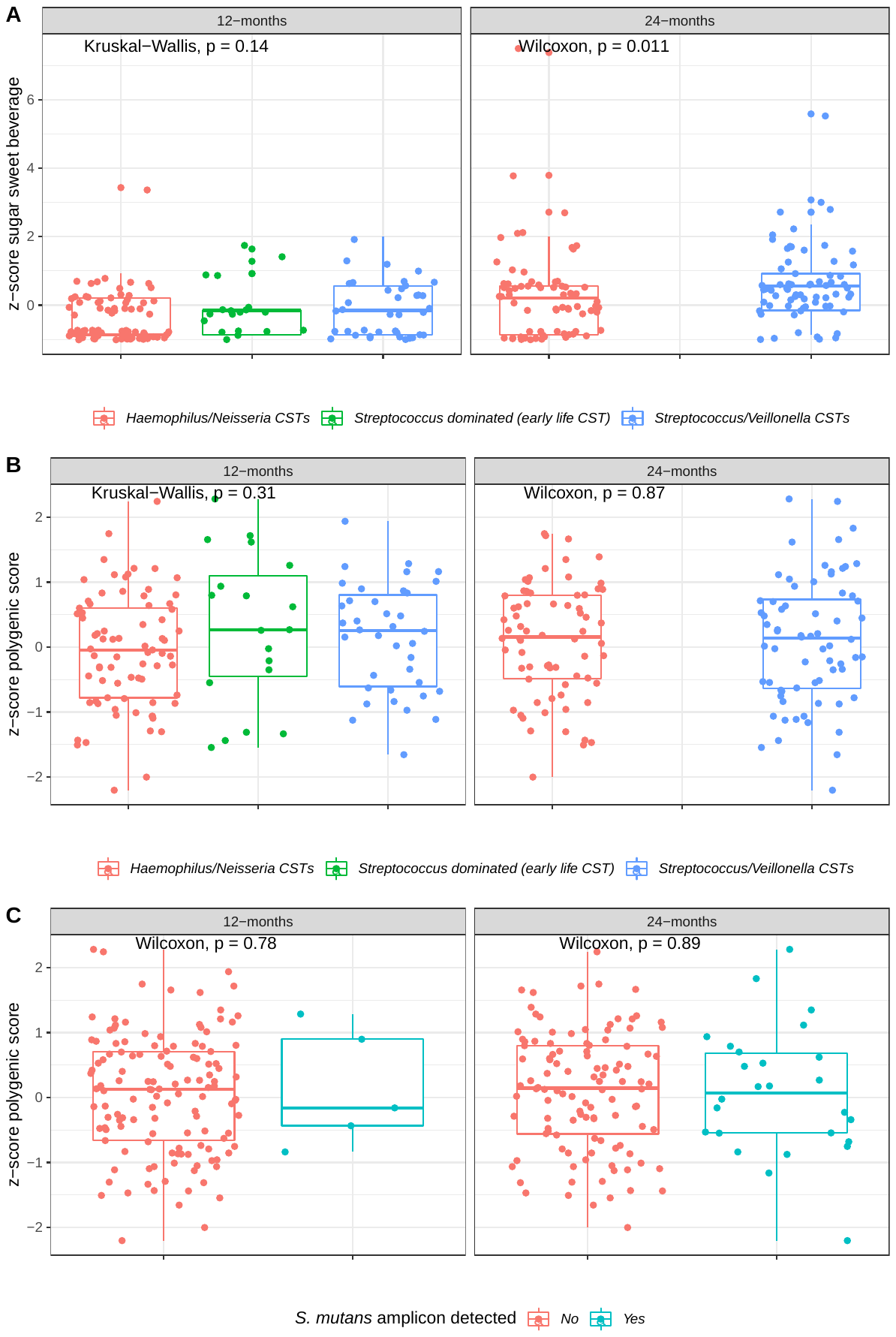

#### Appendix Figure 8: Correlation between alpha diversity measures of salivary microbiome and z-score standardized polygenic risk score for ECC by visit. Left panels: Chao1 measure of richness or number of bacterial species present. Right panels: Shannon index measure of both richness and evenness of distribution of bacterial species. Top panels: 12-month visit salivary microbiome, Bottom panels: 24-month visit salivary microbiome.

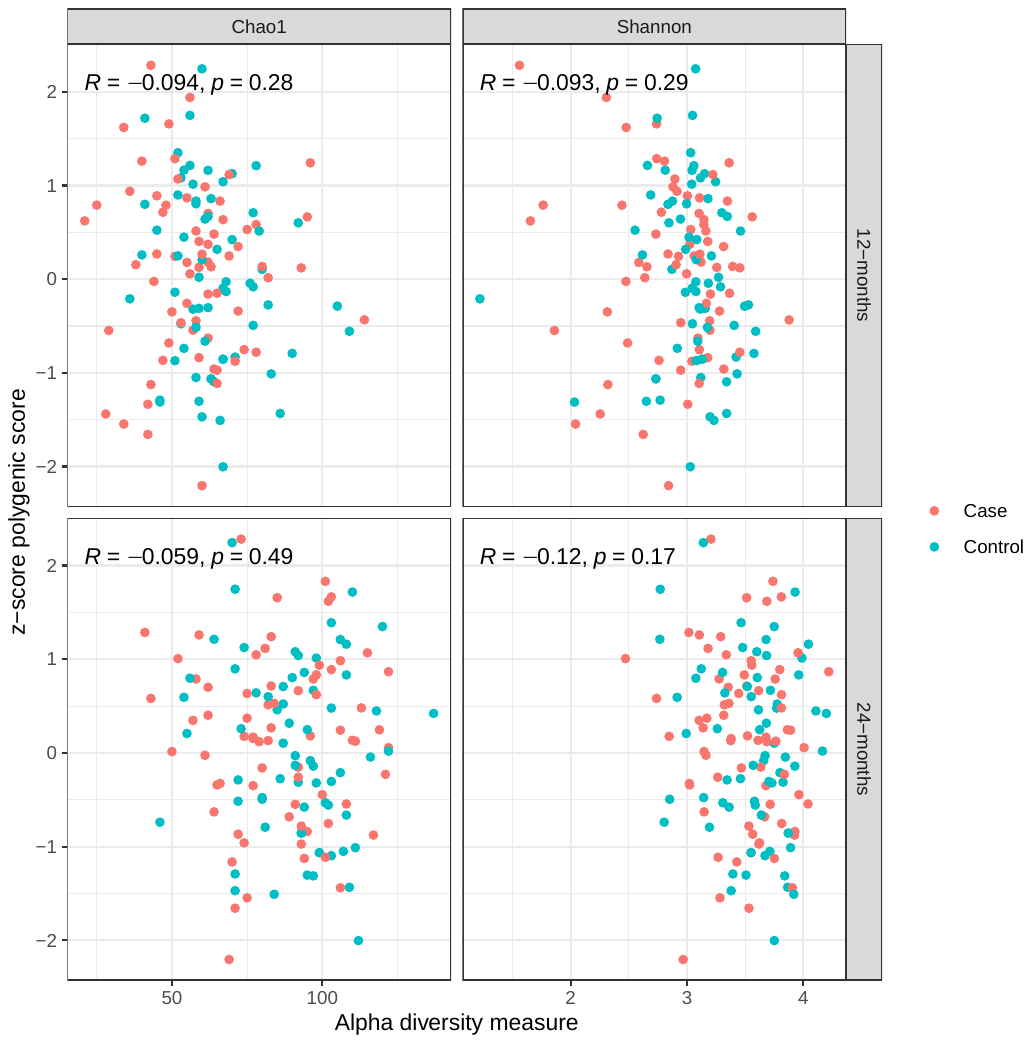

### Appendix Tables

**Appendix Table** **1:** Comparison of included vs excluded sample for analysis of entire longitudinal cohort among 1131 Appalachian children

| Characteristic | Excluded, N = 348^1^ | Included, N = 783^1^ | p-value^2^ |
| --- | --- | --- | --- |
| **Current diagnosis of d1ft** | 33 (13%) | 162 (21%) | 0.008 |
| Unknown | 97 | 0 |  |
| **Age at diagnosis of any primary tooth decay** | 3.02 (2.09, 3.58) | 3.71 (2.99, 4.26) | 0.001 |
| Unknown | 315 | 621 |  |
| **Child age (years)** | 1.98 (0.96, 3.07) | 3.95 (2.09, 4.99) | <0.001 |
| **Child race** |  |  | <0.001 |
| Bi- or Multi-racial | 149 (58%) | 0 (0%) |  |
| White | 110 (42%) | 783 (100%) |  |
| Unknown | 89 | 0 |  |
| **Polygenic score, dichotomous** |  |  | 0.3 |
| Low PGS, <0 | 96 (45%) | 379 (48%) |  |
| High PGS, >=0 | 119 (55%) | 404 (52%) |  |
| Unknown | 133 | 0 |  |
| **Child sex** |  |  | 0.4 |
| Female | 157 (45%) | 373 (48%) |  |
| Male | 191 (55%) | 410 (52%) |  |
| **Child ethnicity** |  |  | 0.064 |
| Not Hispanic/Latino | 313 (93%) | 750 (96%) |  |
| Hispanic/Latino | 23 (6.8%) | 33 (4.2%) |  |
| Unknown | 12 | 0 |  |
| **Maternal education** |  |  | <0.001 |
| Greater than high school education | 236 (68%) | 643 (82%) |  |
| High school or less | 111 (32%) | 140 (18%) |  |
| Unknown | 1 | 0 |  |
| **Maternal age (prenatal visit, years)** | 27.0 (23.0, 32.0) | 29.0 (26.0, 32.0) | <0.001 |
| **Firstborn child** | 146 (42%) | 394 (50%) | 0.013 |
| Unknown | 3 | 0 |  |
| **Prenatal illness** |  |  | 0.10 |
| Over 1 month before prenatal visit | 316 (93%) | 706 (90%) |  |
| Within 1 month of prenatal visit | 23 (6.8%) | 77 (9.8%) |  |
| Unknown | 9 | 0 |  |
| **Prenatal antibiotic use** | 108 (32%) | 217 (28%) | 0.2 |
| Unknown | 6 | 0 |  |
| **Prenatal smoking** | 147 (43%) | 195 (25%) | <0.001 |
| Unknown | 5 | 0 |  |
| **Breastfeeding duration (years)** | 0.17 (0.00, 0.79) | 0.50 (0.06, 1.21) | <0.001 |
| Unknown | 12 | 0 |  |
| **Ever breastfed** |  |  | <0.001 |
| Ever breastfed | 243 (72%) | 664 (85%) |  |
| Never breastfed | 93 (28%) | 119 (15%) |  |
| Unknown | 12 | 0 |  |
| **z-score PC1** | 0.31 (-0.22, 0.79) | 0.02 (-0.62, 0.68) | 0.011 |
| Unknown | 278 | 0 |  |
| **z-score PC2** | -0.10 (-0.95, 0.85) | -0.04 (-0.65, 0.71) | 0.8 |
| Unknown | 278 | 0 |  |
| **z-score PC3** | -0.35 (-0.70, 0.05) | -0.20 (-0.67, 0.36) | 0.056 |
| Unknown | 278 | 0 |  |
| **z-score PC4** | 0.03 (-0.69, 0.64) | -0.07 (-0.72, 0.72) | 0.8 |
| Unknown | 278 | 0 |  |
| **z-score PC5** | 0.04 (-0.68, 0.78) | -0.02 (-0.67, 0.68) | 0.3 |
| Unknown | 278 | 0 |  |
| ^1^n (%); Median (IQR) | | | |
| ^2^Pearson's Chi-squared test; Wilcoxon rank sum test | | | |

**Appendix Table** **2:** Number of cases (by age-at-diagnosis) and controls included in the complete case effect-modification analysis (n=148) compared to number of children ever diagnosed (by age-at-diagnosis) and number never diagnosed in complete case cumulative genetic risk analysis among 783 Appalachian children

| **Age at primary tooth decay diagnosis** | **n in analysis of case control subset** | **n in analysis of cohort** | **percent represented in case control subset (%)** |
| --- | --- | --- | --- |
| 12-month visit | 1 | 3 | 33 |
| 24-month visit | 14 | 22 | 64 |
| 36-month visit | 35 | 56 | 62 |
| 48-month visit | 22 | 46 | 48 |
| 60-month visit | 5 | 36 | 14 |
| Sampled as control (case control subset) / Never dft (cohort) | 65 | 622 | 10 |
| Note: 6 children were selected in an incidence-density risk set as controls but were later diagnosed with ECC. 2 children were selected into >1 risk set and are represented twice in this table | | | |

**Appendix Table** **3:** Bivariate table of dichotomized z-score standardized polygenic score (PGS) for early childhood caries (ECC) by potential confounders, precision variables and other variables of interest among 783 Appalachian children

| Characteristic | N | Low PGS, <0, N = 379^1^ | High PGS, >=0, N = 404^1^ | p-value^2^ |
| --- | --- | --- | --- | --- |
| **Current diagnosis of d1ft** | 783 | 75 (20%) | 87 (22%) | 0.5 |
| **Child sex** | 783 |  |  | 0.8 |
| Female |  | 182 (48%) | 191 (47%) |  |
| Male |  | 197 (52%) | 213 (53%) |  |
| **Child ethnicity** | 783 |  |  | 0.5 |
| Not Hispanic/Latino |  | 365 (96%) | 385 (95%) |  |
| Hispanic/Latino |  | 14 (3.7%) | 19 (4.7%) |  |
| **Maternal education** | 783 |  |  | 0.7 |
| Greater than high school education |  | 309 (82%) | 334 (83%) |  |
| High school or less |  | 70 (18%) | 70 (17%) |  |
| **Maternal age (prenatal visit, years)** | 783 | 29.1 (5.2) | 29.1 (4.9) | 0.6 |
| **Firstborn child** | 783 | 196 (52%) | 198 (49%) | 0.4 |
| **z-score PC1** | 783 | 0.01 (0.95) | 0.03 (1.07) | >0.9 |
| **z-score PC2** | 783 | 0.04 (1.01) | -0.08 (0.99) | 0.10 |
| **z-score PC3** | 783 | -0.09 (0.92) | 0.05 (1.07) | 0.14 |
| **z-score PC4** | 783 | -0.01 (1.00) | 0.01 (1.01) | 0.8 |
| **z-score PC5** | 783 | -0.03 (1.01) | 0.00 (0.97) | 0.5 |
| **Prenatal illness** | 783 |  |  | 0.5 |
| Over 1 month before prenatal visit |  | 339 (89%) | 367 (91%) |  |
| Within 1 month of prenatal visit |  | 40 (11%) | 37 (9.2%) |  |
| **Prenatal antibiotic use** | 783 | 110 (29%) | 107 (26%) | 0.4 |
| **Prenatal smoking** | 783 | 90 (24%) | 105 (26%) | 0.5 |
| **Breastfeeding duration (years)** | 783 | 0.72 (0.81) | 0.81 (0.91) | 0.3 |
| **Ever breastfed** | 783 |  |  | 0.4 |
| Ever breastfed |  | 317 (84%) | 347 (86%) |  |
| Never breastfed |  | 62 (16%) | 57 (14%) |  |
| **Breastfeeding duration** | 783 |  |  | 0.6 |
| >12 months and <= 24 months |  | 98 (26%) | 102 (25%) |  |
| >24 months |  | 22 (5.8%) | 35 (8.7%) |  |
| >6 months & <= 12 months |  | 59 (16%) | 66 (16%) |  |
| 6 months or less |  | 138 (36%) | 144 (36%) |  |
| Never |  | 62 (16%) | 57 (14%) |  |
| ^1^n (%); Mean (SD) | | | | |
| ^2^Pearson's Chi-squared test; Wilcoxon rank sum test | | | | |

**Appendix Table** **4:** Regression model for any versus no early childhood caries (ECC) and dichotomized z-score standardized polygenic score (PGS) at the 36-, 48-, and 60-month visit. Robust standard errors reported in confidence intervals

|  | **3-year visit** | | | **4-year visit** | | | **5-year visit** | | |
| --- | --- | --- | --- | --- | --- | --- | --- | --- | --- |
|  | ***PR*** | ***CI*** | ***P val*** | ***PR*** | ***CI*** | ***P val*** | ***PR*** | ***CI*** | ***P val*** |
| Base model | 1.5 | 0.95, 2.34 | 0.08 | 1.31 | 0.93, 1.85 | 0.12 | 1.23 | 0.88, 1.72 | 0.22 |
| Demographic model | 1.55 | 0.99, 2.42 | 0.05 | 1.41 | 1.01, 1.96 | 0.04 | 1.26 | 0.91, 1.74 | 0.17 |
| Health behaviors model | 1.37 | 0.88, 2.13 | 0.16 | 1.38 | 1, 1.92 | 0.05 | 1.22 | 0.88, 1.69 | 0.24 |
| Base model: adjusted for z-score standardized genetic ancestry principal components,   Demographic model: base model + child sex + maternal education + maternal age at prenatal visit (years), Health behaviours model: demographic model + prenatal illness + prenatal antibiotics + prenatal smoking + ever breastfed, PR: prevalence ratio, CI: 95% Wald confidence interval | | | | | | | | | |

**Appendix Table** **5:** Comparison of sample included in nested case-control subset (n=148 cases and controls, n=146 unique children) to entire cohort (n=783) among Appalachian children

| Characteristic | In genetic models, N = 637^1^ | In GxE models, N = 146^1^ | p-value^2^ |
| --- | --- | --- | --- |
| **Current diagnosis of d1ft** | 80 (13%) | 82 (56%) | <0.001 |
| **Age at diagnosis of any primary tooth decay** | 4.01 (3.00, 4.97) | 3.12 (2.97, 4.01) | 0.002 |
| *Unknown* | 557 | 64 |  |
| **Polygenic score, dichotomous** |  |  | 0.3 |
| *Low PGS, <0* | 314 (49%) | 65 (45%) |  |
| *High PGS, >=0* | 323 (51%) | 81 (55%) |  |
| **Child sex** |  |  | 0.8 |
| *Female* | 302 (47%) | 71 (49%) |  |
| *Male* | 335 (53%) | 75 (51%) |  |
| **Child ethnicity** |  |  | 0.7 |
| *Not Hispanic/Latino* | 611 (96%) | 139 (95%) |  |
| *Hispanic/Latino* | 26 (4.1%) | 7 (4.8%) |  |
| **Maternal education** |  |  | 0.009 |
| *Greater than high school education* | 534 (84%) | 109 (75%) |  |
| *High school or less* | 103 (16%) | 37 (25%) |  |
| **Maternal age (prenatal visit, years)** | 30.0 (26.0, 32.0) | 29.0 (24.0, 32.0) | 0.10 |
| **Firstborn child** | 328 (51%) | 66 (45%) | 0.2 |
| **Prenatal illness** |  |  | 0.6 |
| *Over 1 month before prenatal visit* | 576 (90%) | 130 (89%) |  |
| *Within 1 month of prenatal visit* | 61 (9.6%) | 16 (11%) |  |
| **Prenatal antibiotic use** | 172 (27%) | 45 (31%) | 0.4 |
| **Prenatal smoking** | 143 (22%) | 52 (36%) | <0.001 |
| **Breastfeeding duration (years)** | 0.54 (0.06, 1.24) | 0.33 (0.03, 1.15) | 0.074 |
| **Ever breastfed** |  |  | 0.082 |
| *Ever breastfed* | 547 (86%) | 117 (80%) |  |
| *Never breastfed* | 90 (14%) | 29 (20%) |  |
| **z-score PC1** | -0.03 (-0.63, 0.59) | 0.33 (-0.45, 0.88) | 0.008 |
| **z-score PC2** | -0.03 (-0.64, 0.71) | -0.09 (-0.69, 0.66) | 0.9 |
| **z-score PC3** | -0.19 (-0.67, 0.38) | -0.23 (-0.70, 0.26) | 0.2 |
| **z-score PC4** | -0.06 (-0.73, 0.67) | -0.08 (-0.65, 0.85) | 0.6 |
| **z-score PC5** | -0.07 (-0.65, 0.67) | 0.12 (-0.68, 0.68) | 0.3 |
| ^1^n (%); Median (IQR) | | | |
| ^2^Pearson's Chi-squared test; Wilcoxon rank sum test | | | |

**Appendix Table** **6:** Comparison of children selected as cases and those selected as controls among 148 Appalachian cases and controls (n=146 unique children) by time-invariate characteristics

| Characteristic | N | Case, N = 77^1^ | Control, N = 71^1^ | p-value^2^ |
| --- | --- | --- | --- | --- |
| **Polygenic score, dichotomous** | 148 |  |  | 0.2 |
| *Low PGS, <0* |  | 31 (40%) | 36 (51%) |  |
| *High PGS, >=0* |  | 46 (60%) | 35 (49%) |  |
| **Polygenic score, z-score** | 148 | 0.13 (0.92) | 0.05 (0.92) | 0.6 |
| **Child sex** | 148 |  |  | 0.8 |
| *Female* |  | 36 (47%) | 35 (49%) |  |
| *Male* |  | 41 (53%) | 36 (51%) |  |
| **Child ethnicity** | 148 |  |  | 0.3 |
| *Not Hispanic/Latino* |  | 75 (97%) | 66 (93%) |  |
| *Hispanic/Latino* |  | 2 (2.6%) | 5 (7.0%) |  |
| **Maternal education** | 148 |  |  | 0.010 |
| *Greater than high school education* |  | 51 (66%) | 60 (85%) |  |
| *High school or less* |  | 26 (34%) | 11 (15%) |  |
| **Maternal age (prenatal visit, years)** | 148 | 27.7 (5.9) | 29.6 (5.1) | 0.049 |
| **Firstborn child** | 148 | 34 (44%) | 32 (45%) | >0.9 |
| **z-score PC1** | 148 | 0.17 (0.96) | 0.24 (1.15) | 0.8 |
| **z-score PC2** | 148 | -0.05 (1.03) | 0.01 (1.09) | 0.8 |
| **z-score PC3** | 148 | -0.16 (0.94) | 0.03 (1.16) | 0.2 |
| **z-score PC4** | 148 | 0.08 (0.95) | 0.00 (0.94) | 0.8 |
| **z-score PC5** | 148 | 0.15 (0.86) | 0.04 (1.14) | 0.5 |
| **Prenatal illness** | 148 |  |  | 0.2 |
| *Over 1 month before prenatal visit* |  | 66 (86%) | 66 (93%) |  |
| *Within 1 month of prenatal visit* |  | 11 (14%) | 5 (7.0%) |  |
| **Prenatal antibiotic use** | 148 | 26 (34%) | 19 (27%) | 0.4 |
| **Prenatal smoking** | 148 | 34 (44%) | 18 (25%) | 0.017 |
| **Breastfeeding duration (years)** | 148 | 0.66 (0.94) | 0.74 (0.92) | 0.3 |
| **Ever breastfed** | 148 |  |  | 0.2 |
| *Ever breastfed* |  | 58 (75%) | 60 (85%) |  |
| *Never breastfed* |  | 19 (25%) | 11 (15%) |  |
| ^1^n (%); Mean (SD) | | | | |
| ^2^Pearson's Chi-squared test; Wilcoxon rank sum test; Fisher's exact test | | | | |

**Appendix Table** **7:** Comparison of children selected as cases and those selected as controls among 148 Appalachian cases and controls (n= 146 unique children) by time-invariate characteristics

|  | 1-year visit | | | 2-year visit | | |
| --- | --- | --- | --- | --- | --- | --- |
| Characteristic | Case, N = 69^1^ | Control, N = 65^1^ | p-value^2^ | Case, N = 72^1^ | Control, N = 66^1^ | p-value^2^ |
| **Polygenic score, dichotomous** |  |  | 0.2 |  |  | 0.2 |
| *Low PGS, <0* | 28 (41%) | 34 (52%) |  | 28 (39%) | 33 (50%) |  |
| *High PGS, >=0* | 41 (59%) | 31 (48%) |  | 44 (61%) | 33 (50%) |  |
| **Polygenic score, z-score** | 0.08 (0.91) | 0.02 (0.93) | 0.7 | 0.13 (0.91) | 0.05 (0.93) | 0.6 |
| **Bacterial community state types (CST)** |  |  | <0.001 |  |  | <0.001 |
| *Haemophilus/Neisseria CSTs* | 25 (36%) | 52 (80%) |  | 21 (29%) | 49 (74%) |  |
| *Streptococcus dominated (early life CST)* | 14 (20%) | 5 (7.7%) |  |  |  |  |
| *Streptococcus/Veillonella CSTs* | 30 (43%) | 8 (12%) |  | 51 (71%) | 17 (26%) |  |
| **z-score sugar-sweetened beverage consumption** | -0.05 (0.77) | -0.48 (0.65) | <0.001 | 0.75 (1.34) | 0.10 (0.95) | <0.001 |
| **Currently breastfed** |  |  | 0.5 |  |  | 0.9 |
| *Currently breastfed* | 18 (26%) | 20 (31%) |  | 6 (8.3%) | 5 (7.6%) |  |
| *Not currently breastfed* | 51 (74%) | 45 (69%) |  | 66 (92%) | 61 (92%) |  |
| **Number of primary teeth present** | 6.3 (2.7) | 6.3 (2.6) | >0.9 | 16.53 (1.91) | 16.14 (1.70) | 0.2 |
| **Child antibiotics within 3 months of visit** | 19 (28%) | 17 (26%) | 0.9 | 19 (26%) | 20 (30%) | 0.6 |
| **Streptococcus mutans amplicon detected in saliva sample** | 4 (5.8%) | 1 (1.5%) | 0.4 | 24 (33%) | 2 (3.0%) | <0.001 |
| **Diagnosis/matching visit** |  |  | 0.7 |  |  | 0.8 |
| *36-month visit* | 32 (46%) | 29 (45%) |  | 32 (44%) | 30 (45%) |  |
| *12-month visit* | 1 (1.4%) | 3 (4.6%) |  | 0 (0%) | 0 (0%) |  |
| *24-month visit* | 12 (17%) | 13 (20%) |  | 13 (18%) | 13 (20%) |  |
| *48-month visit* | 19 (28%) | 13 (20%) |  | 22 (31%) | 16 (24%) |  |
| *60-month visit* | 5 (7.2%) | 7 (11%) |  | 5 (6.9%) | 7 (11%) |  |
| ^1^n (%); Mean (SD) | | | | | | |
| ^2^Pearson's Chi-squared test; Wilcoxon rank sum test; Fisher's exact test | | | | | | |

**Appendix Table** **8:** Regression results from regression of case status against 12-month salivary microbial community state type (CST) and z-score standardized polygenic score dichotomized at 0 (PGS) among 138 Appalachian cases and controls (n=136 unique children)

|  |  |  | Base model | | | Full model | | | Full interaction model | | |
| --- | --- | --- | --- | --- | --- | --- | --- | --- | --- | --- | --- |
| **Characteristic** | **N** | **Event N** | **OR** | **95% CI** | **p-value** | **OR** | **95% CI** | **p-value** | **OR** | **95% CI** | **p-value** |
| Polygenic score, dichotomous |  |  |  |  |  |  |  |  |  |  |  |
| Low PGS, <0 | 61 | 28 | — | — |  | — | — |  | — | — |  |
| High PGS, >=0 | 77 | 44 | 1.59 | 0.78, 3.26 | 0.2 | 1.87 | 0.79, 4.42 | 0.2 | 4.83 | 1.29, 18.2 | 0.020 |
| Bacterial community state types (CST) |  |  |  |  |  |  |  |  |  |  |  |
| *Haemophilus/Neisseria CSTs* | 70 | 21 |  |  |  | — | — |  | — | — |  |
| *Streptococcus/Veillonella CSTs* | 68 | 51 |  |  |  | 7.48 | 3.06, 18.3 | <0.001 | 23.8 | 5.28, 107 | <0.001 |
| Polygenic score, dichotomous * Bacterial community state types (CST) | 138 | 72 |  |  |  |  |  |  |  |  |  |
| *High PGS, >=0 * Streptococcus/Veillonella CSTs* | 37 | 27 |  |  |  |  |  |  | 0.15 | 0.02, 0.93 | 0.041 |
| OR=Odds ratio, CI=95% confidence interval (Wald), Base model: adjusted for first 5 principal components of genetic ancestry and visit of case diagnosis/control matching. Full model: adjusted for base model covariates + child sex + maternal education + community state type + breast feeding duration (years) + z-score sugar-sweetened beverage consumption + number of primary teeth present + child receipt of antibiotics in past 3-months | | | | | | | | | | | |

**Appendix Table** **9:** Regression results from regression of case status against salivary microbial community state type (CST) and z-score standardized polygenic score dichotomized at 0 (PGS) among 134 Appalachian cases and controls (n=132)

|  |  |  | Base model | | | Full model | | | Full interaction model | | |
| --- | --- | --- | --- | --- | --- | --- | --- | --- | --- | --- | --- |
| **Characteristic** | **N** | **Event N** | **OR** | **95% CI** | **p-val** | **OR** | **95% CI** | **p-val** | **OR** | **95% CI** | **p-val** |
| Polygenic score, dichotomous |  |  |  |  |  |  |  |  |  |  |  |
| Low PGS, <0 | 62 | 28 | — | — |  | — | — |  | — | — |  |
| High PGS, >=0 | 72 | 41 | 1.60 | 0.78, 3.26 | 0.2 | 1.20 | 0.51, 2.84 | 0.7 | 2.48 | 0.83, 7.41 | 0.10 |
| Bacterial community state types (CST) |  |  |  |  |  |  |  |  |  |  |  |
| *Haemophilus/Neisseria CSTs* | 77 | 25 |  |  |  | — | — |  | — | — |  |
| *Streptococcus dominated (early life CST)* | 19 | 14 |  |  |  | 8.92 | 1.96, 40.6 | 0.005 | 19.1 | 2.20, 166 | 0.008 |
| *Streptococcus/Veillonella CSTs* | 38 | 30 |  |  |  | 9.84 | 3.15, 30.7 | <0.001 | 68.5 | 5.86, 800 | <0.001 |
| Polygenic score, dichotomous * Bacterial CST | 134 | 69 |  |  |  |  |  |  |  |  |  |
| *High PGS,>=0*Streptococcus dominated CST* | 11 | 8 |  |  |  |  |  |  | 0.21 | 0.02, 2.72 | 0.2 |
| *High PGS,>=0*Streptococcus/Veillonella CSTs* | 24 | 17 |  |  |  |  |  |  | 0.06 | 0.00, 0.92 | 0.043 |
| OR=Odds ratio, CI=95% confidence interval (Wald), Base model: adjusted for first 5 principal components of genetic ancestry and visit of case diagnosis/control matching. Full model: adjusted for base model covariates + child sex + maternal education + community state type + breast feeding duration (years) + z-score sugar-sweetened beverage consumption + number of primary teeth present + child receipt of antibiotics in past 3-months | | | | | | | | | | | |

**Appendix Table** **10:** Regression results from regression of case status against 24-month Streptococcus mutans amplicon presence and z-score standardized polygenic score dichotomized at 0 (PGS) among Appalachian children with data available at 24-month visit among138 Appalachian cases and controls (n=136 unique children)

|  |  |  | Full model | | | Full interaction model | | |
| --- | --- | --- | --- | --- | --- | --- | --- | --- |
| **Characteristic** | **N** | **Event N** | **OR** | **95% CI** | **p-value** | **OR** | **95% CI** | **p-value** |
| Polygenic score, dichotomous |  |  |  |  |  |  |  |  |
| Low PGS, <0 | 61 | 28 | — | — |  | — | — |  |
| High PGS, >=0 | 77 | 44 | 1.80 | 0.78, 4.17 | 0.2 | 1.91 | 0.80, 4.55 | 0.14 |
| *Streptococcus mutans amplicon detected in saliva sample* |  |  |  |  |  |  |  |  |
| *Polygenic score, dichotomous * Streptococcus mutans amplicon detected in saliva sample* | 138 | 72 |  |  |  |  |  |  |
| High PGS, >=0 * Yes | 13 | 12 |  |  |  | 0.36 | 0.01, 9.54 | 0.5 |
| OR=Odds ratio, CI=95% confidence interval (Wald), Full model: adjusted for base model covariates + child sex + maternal education + S. mutans presence + breast feeding duration (years) + z-score sugar-sweetened beverage consumption + number of primary teeth present + child receipt of antibiotics in past 3-months | | | | | | | | |

**Supplemental Note on Competing Antagonism**

Consider a hypothetical population in which a cariogenic microbiome, *M,* and a host gene, *G,* are independent and sufficient causes for ECC – which is to say that having either *M* or *G* will result in ECC, and that *M* and *G* do not influence the likelihood of the other. Further, let there be some individuals in the population who are **blessed** – i.e., regardless of exposure to *G* and *M*, they will never experience ECC, and some individuals who are **cursed** – i.e., regardless of *G* or *M* exposure, they will always get ECC (perhaps due to other, unknown sufficient causes), and these individuals are evenly dispersed across exposure statuses. Consider the following illustrative scenarios.

Scenario 1) **Hypothetical population of N=100 with 100% exposure to *M*, 10% exposure to *G,* 10% blessed individuals and 10% cursed individuals.**

There will be 10 individuals with *G:*

- 1 is cursed,
- 1 is blessed.
- 10 are exposed to *M*

There will be 90 individuals without *G*:

- 9 are cursed
- 9 are blessed
- 90 are exposed to *M*

We can calculate the risk ratio for ECC given *G.* Since all individuals in the population were exposed to *M*, a sufficient cause for ECC, all non-blessed individuals will have ECC

|  | ECC yes | ECC no | Total |
| --- | --- | --- | --- |
| *G* yes | 9 | 1 | 10 |
| *G* no | 81 | 9 | 90 |

RR= (9/10 / 81/90 ) = 0.9/0.9= 1🡪 *G* has no association with ECC. Even though *G* would have caused ECC in the 9 exposed individuals *had they been unexposed to M*, this is undetectable epidemiologically.

Note that in this population, the effect of *M* is also undetectable, because there is 100% exposure. To observe an effect of *either G or M*, there must be individuals unexposed to *both G and M*, even though *G* and *M* are independent and in fact are not biologically interacting.

Scenario 2) **Hypothetical population of N=100 with 0% exposure to *M*, 10% exposure to *G,* 10% blessed individuals and 10% cursed individuals.**

There will be 10 individuals with *G:*

- 1 is cursed,
- 1 is blessed.
- 0 are exposed to *M*

There will be 90 individuals without *G*:

- 9 are cursed
- 9 are blessed
- 0 are exposed to *M*

We can calculate the risk ratio for ECC given *G.* Since no individuals in the population were exposed to *M*, a sufficient cause for ECC, only cursed individuals and those with *G* will have ECC.

|  | ECC yes | ECC no | Total |
| --- | --- | --- | --- |
| *G* yes | 9 | 1 | 10 |
| *G* no | 9 | 81 | 90 |

RR = 9/10 / 9/90 = 0.9/ 0.1 = 9🡪 *G* has a strong positive association with ECC

Scenario 3) **Hypothetical population of N=100 with 50% exposure to *M*, 10% exposure to *G,* 10% blessed individuals and 10% cursed individuals.**

There will be 10 individuals with *G:*

- 1 is cursed,
- 1 is blessed.
- 5 are exposed to *M*

There will be 90 individuals without *G*:

- 9 are cursed (of whom half, i.e. 4.5 are exposed to *M)*
- 9 are blessed (of whom half, i.e. 4.5 are exposed to *M)*
- 45 are exposed to *M,* and for 36 of them *M* is causal (since there are 9 individuals who were exposed to *M* but were either blessed or cursed)

We can calculate the risk ratio for ECC given *G.* Non-blessed individuals in the population exposed to *M*, a sufficient cause for ECC, or with *G,* as well as cursed individuals will have ECC:

|  | ECC yes | ECC no | Total |
| --- | --- | --- | --- |
| *G* yes | 9 | 1 | 10 |
| *G* no | 9 (cursed) + 36(causal exposure to M) = 45 | 9 (blessed)+ 36 (not exposed to M)=45 | 90 |

RR=9/10 / 45/90 = 0.9/0.5 = 1.8 🡪 G has a positive association with ECC

In this scenario we can also consider the effect of stratifying by *M:*

**Among those *M* yes**

|  | ECC yes | ECC no | Total |
| --- | --- | --- | --- |
| *G* yes | 4.5 | 0.5 (blessed) | 5 |
| *G* no | 4.5 (cursed) + 36(causal exposure to M) = 40.5 | 4.5 (blessed) | 45 |

RR =4.5/5 / 40.5/45 = 1

**Among those *M* no**

|  | ECC yes | ECC no | Total |
| --- | --- | --- | --- |
| *G* yes | 4.5 | 0.5 (blessed) | 5 |
| *G* no | 4.5 (cursed) | 4.5 (blessed) + 36(not exposed to M) =40.5 | 45 |

RR =4.5/5 / 4.5/45 = 9

Since there are now individuals in the population who are unexposed to both *M* and *G*, we can consider an interaction table like the one presented in this manuscript, where the reference group for interaction is those unexposed to both *M* and *G:*

| Exposure status | Total | Blessed | Cursed | With ECC | Risk |
| --- | --- | --- | --- | --- | --- |
| *G* and *M* exposed | 5 | .5 | .5 | 4.5 (all but blessed) | 4.5/5=0.9 |
| *G* exposed and *M* unexposed | 5 | .5 | .5 | 4.5 (all but blessed) | 4.5/5=0.9 |
| *G* unexposed and *M* exposed | 45 | 4.5 | 4.5 | 40.5 (all but blessed) | 40.5/45=0.9 |
| *G* and *M* unexposed | 45 | 4.5 | 4.5 | 4.5 (only cursed) | 4.5/45=0.1 |

|  | *M* unexposed | *M* exposed |
| --- | --- | --- |
| *G* unexposed | 1 (reference) | RR = 0.9/0.1=9 |
| *G* exposed | RR=0.9/0.1=9 | RR=0.9/0.1=9 |

We can calculate a measure of multiplicative interaction as 9/(9*9)=1/9, which is indicative of submultiplicative interaction. Similarly, we can calculate the RERI as 9-9-9+1 = -8, which is indicative of subadditive interaction.

Note that if the idea of half persons is bothersome, we can multiply all cells by 2 and consider a population of 200 instead of 100.

Since the statistical power to detect an association depends on the sample size and the effect size, a larger sample size will be needed to detect the RR of 1.8 in Scenario 3 than the RR of 9 in Scenario 2. This illustrates how the prevalence and strength of an environmental risk factor can influence the ability to detect genetic associations. The example of stratification by *M* in Scenario 3 illustrates how a genetic risk factor may only be detectable among those without the strong environmental risk factor.

Keep in mind that in Scenario 1 the causal effect of *G* is indeed null: under the hypothetical situation in which we genetically edit *G* to no longer exist, there would be no change in the population incidence of ECC because all individuals would still be exposed to *M*. However, if we were able to entirely prevent *M* (scenario 2), *G* would have a causal effect. The existence of such statistical interactions, even in the absence of any biological interaction, can still be meaningful for prioritizing interventions and identifying susceptible populations. In a situation with limited resources, we might decide to prioritize intervening on *M,* which is responsible for more cases. When resources are unlimited or in populations where *M* is less prevalent, we might consider staged preventative recommendations: i.e., all children with *M* or *G* should receive more dental examinations a year, than those without *M* and *G.*

1.	Haworth S, Shungin D, van der Tas JT, et al (2018) [Consortium-based genome-wide meta-analysis for childhood dental caries traits](https://doi.org/10.1093/hmg/ddy237). Human Molecular Genetics 27:3113–3127

2.	Mansournia MA, Jewell NP, Greenland S (2017) [Casecontrol matching: effects, misconceptions, and recommendations](https://doi.org/10.1007/s10654-017-0325-0). European Journal of Epidemiology 33:5–14

3.	Kozich JJ, Westcott SL, Baxter NT, Highlander SK, Schloss PD (2013) [Development of a Dual-Index Sequencing Strategy and Curation Pipeline for Analyzing Amplicon Sequence Data on the MiSeq Illumina Sequencing Platform](https://doi.org/10.1128/aem.01043-13). Applied and Environmental Microbiology 79:5112–5120

4.	Callahan BJ, McMurdie PJ, Rosen MJ, Han AW, Johnson AJA, Holmes SP (2016) [DADA2: High-resolution sample inference from Illumina amplicon data](https://doi.org/10.1038/nmeth.3869). Nature Methods 13:581–583

5.	Chen T, Yu W-H, Izard J, Baranova OV, Lakshmanan A, Dewhirst FE (2010) [The Human Oral Microbiome Database: a web accessible resource for investigating oral microbe taxonomic and genomic information](https://doi.org/10.1093/database/baq013). Database 2010:baq013–baq013

6.	Davis NM, Proctor DM, Holmes SP, Relman DA, Callahan BJ (2018) Simple statistical identification and removal of contaminant sequences in marker-gene and metagenomics data. Microbiome. <https://doi.org/10.1186/s40168-018-0605-2>

7.	Robinson D, Hayes A, Couch S (2022) [Broom: Convert statistical objects into tidy tibbles](https://CRAN.R-project.org/package=broom).

8.	Wickham H (2021) [Tidyverse: Easily install and load the tidyverse](https://CRAN.R-project.org/package=tidyverse).

9.	Wickham H, François R, Henry L, Müller K (2022) [Dplyr: A grammar of data manipulation](https://CRAN.R-project.org/package=dplyr).

10.	Wickham H (2022) [Stringr: Simple, consistent wrappers for common string operations](https://CRAN.R-project.org/package=stringr).

11.	Lüdecke D (2021) [Sjlabelled: Labelled data utility functions](https://strengejacke.github.io/sjlabelled/).

12.	Ripley B (2021) [MASS: Support functions and datasets for venables and ripley’s MASS](http://www.stats.ox.ac.uk/pub/MASS4/).

13.	Oksanen J, Blanchet FG, Friendly M, et al (2020) [Vegan: Community ecology package](https://CRAN.R-project.org/package=vegan).

14.	Morgan M (2020) DirichletMultinomial: Dirichlet-multinomial mixture model machine learning for microbiome data.

15.	Sjoberg DD, Curry M, Larmarange J, Lavery J, Whiting K, Zabor EC (2022) [Gtsummary: Presentation-ready data summary and analytic result tables](https://CRAN.R-project.org/package=gtsummary).

16.	Hugh-Jones D (2022) [Huxtable: Easily create and style tables for LaTeX, HTML and other formats](https://hughjonesd.github.io/huxtable/).

17.	Gohel D, Skintzos P (2022) [Flextable: Functions for tabular reporting](https://CRAN.R-project.org/package=flextable).

18.	Wickham H, Chang W, Henry L, Pedersen TL, Takahashi K, Wilke C, Woo K, Yutani H, Dunnington D (2022) [ggplot2: Create elegant data visualisations using the grammar of graphics](https://CRAN.R-project.org/package=ggplot2).

19.	Kassambara A (2020) [Ggpubr: ggplot2 based publication ready plots](https://rpkgs.datanovia.com/ggpubr/).
